# Spatial proteomics identifies a myeloid-dominated, compartmentalized microenvironment and a proliferation-associated prognostic signature in clear cell sarcoma

**DOI:** 10.64898/2026.11.20.26368290

**Authors:** Simon Haefliger, Nikolaus Deigendesch, Benjamin Ehret, Lasse Meyer, Michelle Daniel, Christopher Davies, Akanksha Farswan, Agnieszka Wozniak, Patrick Schöffski, Natalie de Souza, Antoine Italiano, Daniel Baumhoer, Chantal Pauli, Felix Meissner, Stephane Chevrier, Nischalan Pillay, Bernd Bodenmiller, Clear Cell Sarcoma Consortium

**Affiliations:** Department of Quantitative Biomedicine, University of Zürich, Zürich, Switzerland; Institute for Molecular Health Sciences, ETH Zürich, Zürich, Switzerland; Life Science Zürich Graduate School, ETH and University of Zürich, Zürich, Switzerland; Institute of Medical Genetics and Pathology, University Hospital Basel, University of Basel, Basel, Switzerland; Navignostics AG, Horgen, Switzerland; Institute of Innate Immunity, Department of Systems Immunology and Proteomics, University of Bonn, Medical Faculty, Bonn, Germany; Research Department of Pathology, Cancer Institute, University College London (UCL), London, UK; Department of General Medical Oncology, University Hospitals Leuven, and Laboratory of Experimental Oncology, KU Leuven, Leuven Cancer Institute, Leuven, Belgium; Institute of Molecular Systems Biology, Department of Biology, ETH Zürich, Zürich, Switzerland; DITEP, Gustave Roussy, Villejuif, France; HOCH Health Ostschweiz, Institute of Pathology, Reference Center for Bone and Craniofacial Tumors, St. Gallen, Switzerland; Institute of Surgical Pathology, University Hospital Zürich, Zürich, Switzerland; Department of Cellular and Molecular Pathology, Royal National Orthopaedic Hospital (RNOH) NHS Trust, Stanmore, Middlesex, UK

**Keywords:** Clear cell sarcoma, sarcoma, imaging mass cytometry, deep visual proteomics, spatial proteomics, tumour microenvironment, antibody–drug conjugates

## Abstract

Clear cell sarcoma (CCS) is a rare soft-tissue sarcoma with poor prognosis and limited therapeutic options. Its tumour microenvironment and tumour cell heterogeneity remain largely unexplored. Within the Clear Cell Sarcoma Consortium, we used imaging mass cytometry with two antibody panels to map more than 5 million cells from a multi-institutional, clinically annotated cohort of 52 patients. CCS was immune-cold: myeloid cells, principally CD68+ and CD163+ macrophages, dominated the tumour centre, whereas the invasive front contained a more diverse immune infiltrate. A proliferative tumour cell phenotype was associated with shorter overall survival. Deep visual proteomics identified an associated proteomic signature, and deep-learning models predicted the phenotype from haematoxylin-and-eosin images. c-MET and B7-H3 emerged as candidate antibody–drug conjugate targets with broadly homogeneous expression. Together, this work provides a detailed map of the spatial proteomic landscape of CCS and highlights candidate markers for patient stratification and potential therapeutic targets.

**STATEMENT OF SIGNIFICANCE:** Clear cell sarcoma is a rare malignancy with few treatment options. Our spatial single-cell proteomic atlas reveals a myeloid-dominated, compartmentalized tumour microenvironment and identifies a proliferative tumour cell state recognizable on routine H&E-stained sections and associated with shorter survival. It also nominates c-MET and B7-H3 as antibody–drug conjugate targets.

## INTRODUCTION

Clear cell sarcoma (CCS) is a rare, translocation-associated soft-tissue sarcoma that predominantly arises in the tendons and aponeuroses of the lower extremities in young adults (Enzinger, 1965; Gonzaga et al., 2018; Kawai et al., 2007). Although it may initially present as an indolent lesion, CCS is clinically aggressive, with early metastatic spread, frequent local recurrence and poor long-term survival; overall survival rates across all stages are approximately 50% at five years and 40% at ten years (Bianchi et al., 2014; Clark et al., 2008; Gonzaga et al., 2018; Wang et al., 2025). Therapeutic options for advanced disease remain extremely limited (Wetterwald et al., 2023). CCS is largely refractory to conventional chemotherapy and radiotherapy, and targeted and immune-based therapies have shown only modest clinical activity (Gronchi et al., 2021; Jones et al., 2011; Martin-Broto et al., 2020; Martin-Broto et al., 2026; Merchant et al., 2016; Schöffski et al., 2017). CCS is classified as an ultra-rare cancer, with a reported annual incidence of fewer than 0.5 cases per million people, which limits clinical trials and the discovery of actionable biological targets (Stacchiotti et al., 2021). CCS tumour tissue samples from multi-institutional cohorts, with linked clinical data, are therefore also sparse and particularly valuable for advancing the understanding of this disease. At the molecular level, CCS is defined by recurrent *EWSR1* rearrangements, most commonly *EWSR1::ATF1*, with rarer fusions involving genes encoding other members of the CREB transcription factor family (Straessler et al., 2013; Yamada et al., 2013; Zucman et al., 1993). These oncogenic fusions encode aberrant transcription factors that result in melanocytic differentiation, accounting for melanocytic marker expression and the historical designation of CCS as melanoma of soft parts (Straessler et al., 2013; Yamada et al., 2013; Zucman et al., 1993).

Recent genomic and transcriptomic studies have begun to define the genomic landscape of CCS (Lee et al., 2021; Panza et al., 2021; Rasmussen et al., 2023). However, these studies have relied almost exclusively on bulk profiling approaches, masking cellular heterogeneity and eliminating spatial context. As a result, fundamental questions regarding the composition, spatial organization, and functional interplay of the CCS tumour microenvironment (TME) remain unanswered.

Here, we applied high-dimensional, spatially resolved single-cell proteomic profiling to specimens from a multi-institutional cohort of clinically characterized CCS patients assembled through the Clear Cell Sarcoma Consortium. This retrospective exploratory approach enabled systematic mapping of the tumour and immune compartments in CCS and the identification of prognostic features that might aid patient stratification and the development of individual treatment approaches.

## RESULTS

### Clear cell sarcoma cohort

The aim of the study was to use spatial proteomics to analyse the tumour ecosystem in CCS and to relate it to prognostic features and potential therapeutic targets (**Fig. 1A).** We assembled formalin-fixed paraffin-embedded (FFPE) samples from a clinically annotated cohort of CCS patients for spatial proteomic profiling (**Fig. 1B; Sup. Table S1**). Diagnoses were established according to the 2020 WHO Classification of Soft Tissue and Bone Tumours (WHO Classification of Tumours Editorial Board, 2020) and were confirmed by an expert sarcoma pathologist who reviewed H&E-stained slides of samples from each case. To minimize treatment-related confounding, only specimens obtained before any systemic therapy were included in our analysis. After central case review, 56 patients were retained for imaging mass cytometry (IMC) analysis. Regions of interest (ROIs) were selected by a pathologist and classified as tumour centre, invasive front, or lymph-node metastasis for downstream analyses (**Fig. 1E**). From these ROIs, we constructed three tissue microarrays (TMAs). Laser-microdissected regions from a subset of these patients were additionally profiled by deep visual proteomics (DVP), and H&E-stained sections of the same ROIs were used for deep-learning analysis (**Fig. 1A**; Methods).

**FIGURE 1.**
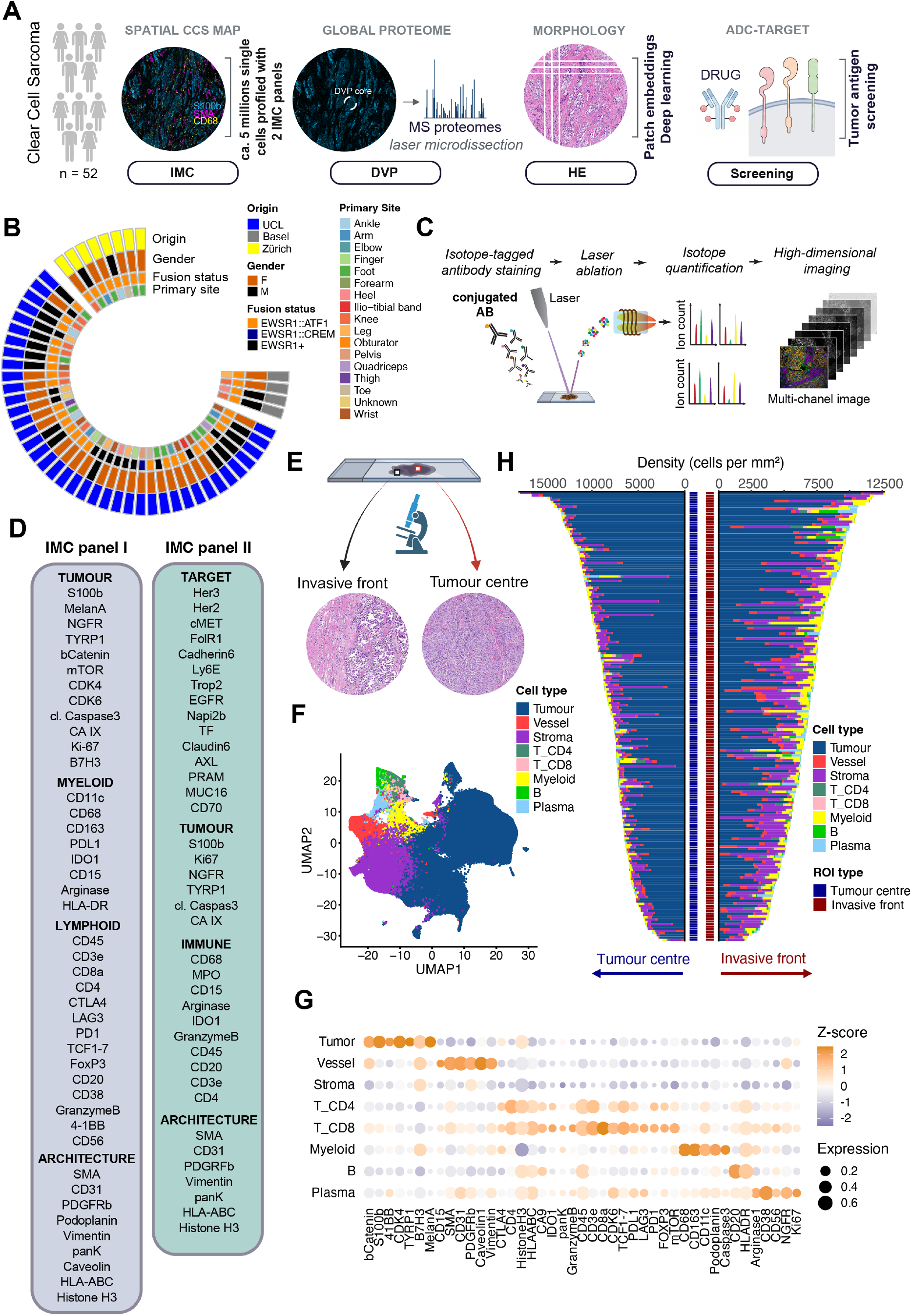
High-dimensional imaging of clear cell sarcoma. **(A)** Overview of the methods used in the study (created with BioRender.com). **(B)** Circos plot of clinical metadata for the 52 patients for whom IMC data were collected. **(C)** Overview of the IMC workflow: TMA sections were stained with metal-tagged antibodies, regions of interest (ROIs) were ablated by laser, and isotope signals were quantified by time-of-flight mass spectrometry to reconstruct multi-channel images (created with BioRender.com). **(D)** Markers targeted by the antibodies of IMC panel I and II. **(E)** ROI selection strategy on H&E sections: tumour centre and invasive front areas were selected for profiling (created with BioRender.com). **(F)** Uniform manifold approximation and projection (UMAP) of single cells (10% downsampled for visualization) coloured by major cell type. **(G)** Dot plot of marker expression across cell types; dot size indicates the mean scaled expression and colour denotes z-scored expression. Expression is arcsinh-transformed with a cofactor of 1 before z-scoring. **(H)** Mirrored bar plots showing cell density (cells/mm^2^) per ROI for tumour centre (left, blue axis label) and invasive front (right, red axis label). Colours indicate cell type, and bar lengths sum to the total density for each ROI.

### A single-cell, high-dimensional spatial atlas of CCS

To characterize phenotypic heterogeneity and the spatial organization of tumour cells relative to the TME, we performed high-dimensional, spatial proteomic profiling using IMC (Giesen et al., 2014) (**Fig. 1C**). TMA sections were stained with two metal-conjugated antibody panels (**Fig. 1D**) designed to characterize both the tumour and the immune microenvironment. Panel I included broad immune-lineage markers, activation, exhaustion and proliferation markers for T cells (TCF1/7, PD-1, CTLA-4, LAG-3, FOXP3, granzyme B, Ki-67) and for myeloid cells (HLA-DR, PD-L1, IDO1, arginase-1, CD11c), together with tumour-state and architectural markers (**Fig. 1D; Sup. Table S2; Fig. S1A**). Panel II was designed to spatially screen tumour-antigen expression and identify candidates for antibody–drug conjugate (ADC) targeting (**Fig. 1D; Sup. Table S3; Fig. S2A**). With panel I, we acquired 460 high-dimensional images, which were segmented into approximately 3 million single cells. Image preprocessing and single-cell segmentation were performed using established IMC analysis pipelines (Windhager et al., 2023) (see Methods). After quality control and exclusion of control cores, we retained 427 images from 52 patients, comprising 2,870,352 cells; four patients were excluded from downstream analysis following quality control. With panel II, 412 images from the same 52 patients, comprising 2,513,554 cells, were retained after quality control. No batch effects were apparent on visual inspection of the uniform manifold approximation and projection (UMAP) embeddings with respect to TMA, sample origin, patient identity, or ROI category (**Fig. S1B, Fig. S2B).**

To define cellular phenotypes, we trained an image-based neural network using the CellSighter framework (Amitay et al., 2023) on our dataset (**Fig. S3A–G;** see Methods). This approach enabled classification into eight broad cell types: B cells, plasma cells, CD4⁺ T cells, CD8⁺ T cells, myeloid cells, tumour cells, vessels, and stromal cells. UMAP visualization revealed clear separation of the major cell types into distinct clusters (**Fig. 1F**), accompanied by coherent expression of canonical lineage and state markers (**Fig. 1G**). Tumour cells constituted the dominant population across most samples (**Fig. 1H**).

This single-cell, spatially resolved proteomic atlas of CCS provides a framework for studying tumour-cell heterogeneity and the organization of the immune microenvironment in this rare malignancy.

### CCS tumours are characterized by a compartmentalized immune environment dominated by myeloid cells

Given the critical role of the immune microenvironment and its spatial organization in cancer (Binnewies et al., 2018; Bruni et al., 2020; de Visser & Joyce, 2023; Quail & Joyce, 2013), we next characterized the spatial organization of immune infiltrates in CCS tumours from the 40 patients in the cohort for whom both tumour centres and invasive fronts were imaged. Immune-cell density was markedly lower in the tumour centre than at the invasive front (**Fig. 2A**), consistent with an “immune-cold” tumour core. Further, the tumour centre had a higher fraction of myeloid cells, even though the absolute myeloid cell density was lower at the tumour centre than at the invasive front (**Fig. 2B**). Lymphoid cells showed the opposite pattern, with both higher fraction and higher density at the invasive front than at the tumour centre (**Fig. 2C**). These patterns were also apparent in a summary of per-ROI composition and mean CD3e, CD20, and CD68 marker expression (**Fig. 2D**). Differential abundance analysis further supported enrichment of myeloid cells in the tumour centre and preferential localization of lymphoid cells to the invasive front (**Fig. 2E**; representative tumour centre ROI in **Fig. 2G**). The invasive front had a more diverse immune infiltrate, as reflected by significantly higher Shannon entropy (as a measure of disorder) of immune cells, relative to the tumour centre (**Fig. 2F**). Thus, the sparse immune infiltrate within CCS tumours was predominantly composed of myeloid cells, consistent with its prior characterization as an immune-low type of sarcoma (Petitprez et al., 2020). In line with this, analyses of H&E-stained sections did not identify any tertiary lymphoid structures in samples from this cohort.

**FIGURE 2.**
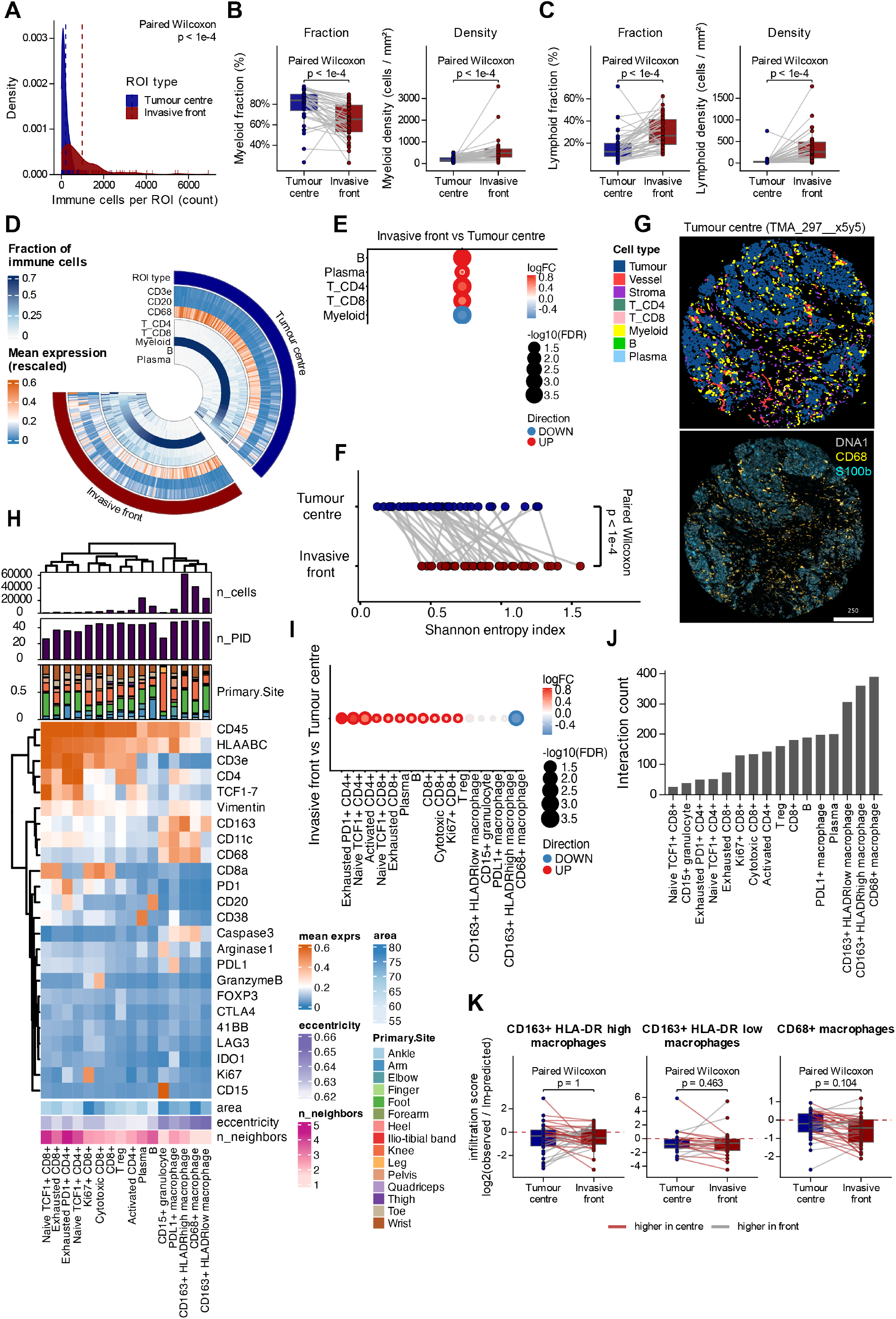
The immune microenvironment of CCS is immune-cold and compartmentalized. **(A)** Distributions of total immune cell counts per ROI, stratified by ROI type for the 40 patients for whom tumour centre and invasive front regions were analysed. P values are indicated (paired Wilcoxon signed-rank test). **(B and C)** Box plots with paired comparisons of percentages and densities (cells/mm²) of (B) myeloid cells and (C) lymphoid cells between tumour centre and invasive front for matched ROIs from the same patients. Points represent individual mean values per patient for the given ROI; grey lines connect paired measurements. Box plots show the median and interquartile range (IQR), and whiskers extend to the most extreme values within 1.5×IQR from the quartiles. P values are indicated (paired Wilcoxon signed-rank test). **(D)** Circular heatmap showing one ROI in each slice, split by region, and showing immune cell type abundance and mean expression of the indicated markers. **(E)** Differential abundance of major immune cell types between invasive front and tumour centre determined by edgeR quasi-likelihood F-test. Dot size indicates −log10 of the false discovery rate (FDR), colour indicates log2 fold change (invasive front vs. tumour centre), and the ring marks cell types called at FDR below 0.1 (Benjamini-Hochberg correction). Red, enriched at the invasive front; blue, enriched in the tumour centre; light grey, not significant. **(F)** Dot plot showing the Shannon entropy of the immune cell-type composition per patient, tumour centre vs. invasive front. P value is indicated (paired Wilcoxon signed-rank test). **(G)** Images of a cell-phenotype map (top) and DNA1, CD68, and S100b marker expression (bottom) for an example tumour centre ROI from patient CCS05. Scale bar, 250 μm. **(H)** Heatmap of marker expression (z-score) per immune subtype. Shown are the total numbers of cells and patients per subtype, the fraction of each subtype per anatomical region, and average spatial single-cell information (area, eccentricity, number of neighbours). Rows and columns are hierarchically clustered using Ward’s method (ward.D2). **(I)** Differential abundance analysis of immune subtypes, as in (E). **(J)** Bar plot ranking tumour interaction partners by interaction count: plotted are the per-ROI mean number of tumour cell neighbours of each indicated cell type, per tumour cell with at least one such neighbour, summed over ROIs. All partner cell types are shown. **(K)** Box plots showing comparison of macrophage infiltration scores between tumour centre and invasive front in paired samples. Each point represents one patient; lines connect matched regional measurements. The infiltration score, log2(observed contact rate / predicted contact rate), is shown for CD163⁺ HLA-DRhigh, CD163⁺ HLA-DRlow and CD68⁺ macrophages respectively. Box plots show the median and IQR, and whiskers extend to the most extreme values within 1.5×IQR from the quartiles. P values are indicated (paired Wilcoxon signed-rank test, Benjamini-Hochberg corrected over the three macrophage populations).

To identify lineage-specific immune states, we applied graph-based clustering to CD8⁺ and CD4⁺ T cells as well as myeloid cell populations, using lineage-specific state markers (**Fig. 2H; Fig. S4A–G).** We identified five CD8⁺ T cell subtypes (cytotoxic CD8⁺, exhausted CD8⁺, Ki-67⁺ CD8⁺, naïve-like TCF1/7⁺ CD8⁺, CD8⁺ T cell without specific marker profile referred to as CD8⁺ T cell), four CD4⁺ T cell subtypes (regulatory T cell (Treg), naïve-like TCF1/7⁺ CD4⁺, activated CD4⁺, and exhausted PD-1⁺ CD4⁺), and five myeloid cell populations (CD15⁺ granulocyte, CD68⁺ macrophage, PD-L1⁺ macrophage, CD163⁺ HLA-DRhigh macrophage, CD163⁺ HLA-DRlow macrophage). Differential abundance analysis revealed an enrichment of CD68⁺ macrophages in the tumour centre, whereas multiple lymphoid states, including naïve-like and exhausted T cells, were enriched at the invasive front (**Fig. 2I**).

Next, we assessed tumour–immune interactions in the tumour centre and invasive front using a neighbourhood-based model that accounts for cell abundance (see Methods). Across both regions, raw tumour–immune interaction counts were dominated by macrophages, with interactions between tumour cells and CD68⁺ macrophages, CD163⁺ HLA-DRhigh, and CD163⁺ HLA-DRlow macrophages together constituting about three-quarters (76%) of all tumour–immune cell contacts (**Fig. 2J; Fig. S4H**). We then focused on these macrophage populations and used an approach that accounted for regional differences in cell abundance to compare infiltration scores between the tumour centre and the invasive front (see Methods and **Fig. S5A–E**). Infiltration scores were comparable between regions for CD163⁺ HLA-DRhigh, CD163⁺ HLA-DRlow and CD68⁺ macrophages (**Fig. 2K** and **Fig. S5A–E**), suggesting similar tumour–macrophage interactions across tumour regions, despite differences in cell density. Deconvolution of bulk transcriptome data (Chen et al., 2018) in a subset of the primary cohort (n = 23 patients) supported a predominantly macrophage-derived myeloid signal in these tumours (**Fig. S4K–M**).

Next, we assessed whether the immune cell composition was associated with patient outcome. In Cox proportional hazards models adjusted for age, sex and metastatic status at diagnosis, no immune cell population was associated with overall survival (OS) at both cut-points (mean and median split); CD15⁺ granulocyte abundance was associated with OS at the mean split only (**Fig. S4I–J**).

For validation of these findings, we performed IMC on an independent CCS cohort from KU Leuven (n = 20 patients passing quality control; **Sup. Table S4**) using the same antibody panel and an analogous analytical approach. Consistent with the primary cohort, this validation cohort exhibited low immune infiltration, dominated by myeloid cells (**Fig. S6A–F**). We also fitted survival models for the immune subtypes in this cohort. However, with only 13 patients with usable follow-up and 10 events, the analysis had limited statistical power, so the lack of a detected association does not provide evidence that no association exists (**Fig. S6K–L**).

CCS was historically referred to as “malignant melanoma of soft parts”. Given the phenotypic similarities between CCS and melanoma, a long-standing question is whether their tumour immune microenvironments also share features. To address this question, we compared the CCS TME with that of cutaneous melanoma (CM), an immunogenic malignancy driven by distinct intrinsic genetic mechanisms and exogenous mutagenic influences of ultraviolet light (**Fig. S7A**) (Akbani et al., 2015; Wetterwald et al., 2023). We analysed a CM cohort (n = 38 patients; 152 ROIs) (**Fig. S7B; Sup. Table S5**), stained with the same IMC antibody panel used for CCS. Using the same phenotyping framework as for CCS, we then jointly analysed immune cells from both cohorts; UMAP analysis showed no cohort-specific segregation suggestive of batch effects (**Fig. S7C**). Compared with CM, CCS showed lower immune cell density and diversity (Shannon entropy), but a higher myeloid fraction (**Fig. S7D–G**). Definition of CD8⁺ T cells, CD4⁺ T cells, and myeloid/macrophage populations across both cohorts (**Fig. S7H**) revealed enrichment of CD68⁺ and CD163⁺ macrophages in CCS upon differential abundance analysis, whereas lymphoid populations, including exhausted PD-1⁺ CD4⁺ and PD-1⁺ CD8⁺ T cells, were enriched in CM (**Fig. S7I**). These macrophages constituted a larger immune cell fraction in CCS despite comparable or lower absolute densities (**Fig. S7J–K**).

Collectively, these findings indicate that CCS tumours are “immune-cold” with a myeloid-dominated, spatially compartmentalized microenvironment and a more immune-diverse invasive front compared with the tumour centre. Despite shared melanocytic differentiation, CCS and CM have distinct immune ecosystems, with greater immune cell abundance and diversity in CM, further supporting the classification of CCS as “immune-cold”.

### A proliferative tumour cell state is associated with CCS patient outcome

Next, we focused on the tumour compartment to determine whether distinct tumour cell phenotypes have prognostic relevance in CCS. We identified 34 tumour clusters via graph-based clustering of tumour cells using markers of cell state (**Fig. 3A**), and subsequently metaclustered these based on mean marker expression into 5 tumour metaclusters (MCs) (**Fig. 3A, Fig. S8A–C).** These tumour MCs separated distinctly on the UMAP embedding (**Fig. 3B**) and were further evaluated by visual inspection of the corresponding IMC images (**Fig. S8D**). Each MC displayed a distinct expression pattern with specific dominant markers: MC1 was characterized by NGFR expression, MC3 by Ki-67, MC4 by HLA-DR and MC5 by TYRP1; only MC2 lacked a defining marker-expression pattern from our antibody panel (**Fig. 3A, Fig. S8D**). All tumour MCs were present both in the tumour centre and at the invasive front, and most ROIs were dominated by one to three MCs (**Fig. 3C**). Shannon entropy was higher at the invasive front than in the tumour centre, indicating greater phenotypic diversity at the tumour margin (**Fig. 3D**). However, no MC was significantly enriched at one location versus the other after correction for multiple testing (**Fig. 3E; Fig. S8H**). In contrast to the immune infiltrate, therefore, tumour MCs in CCS do not exhibit spatial compartmentalization of their composition.

**FIGURE 3.**
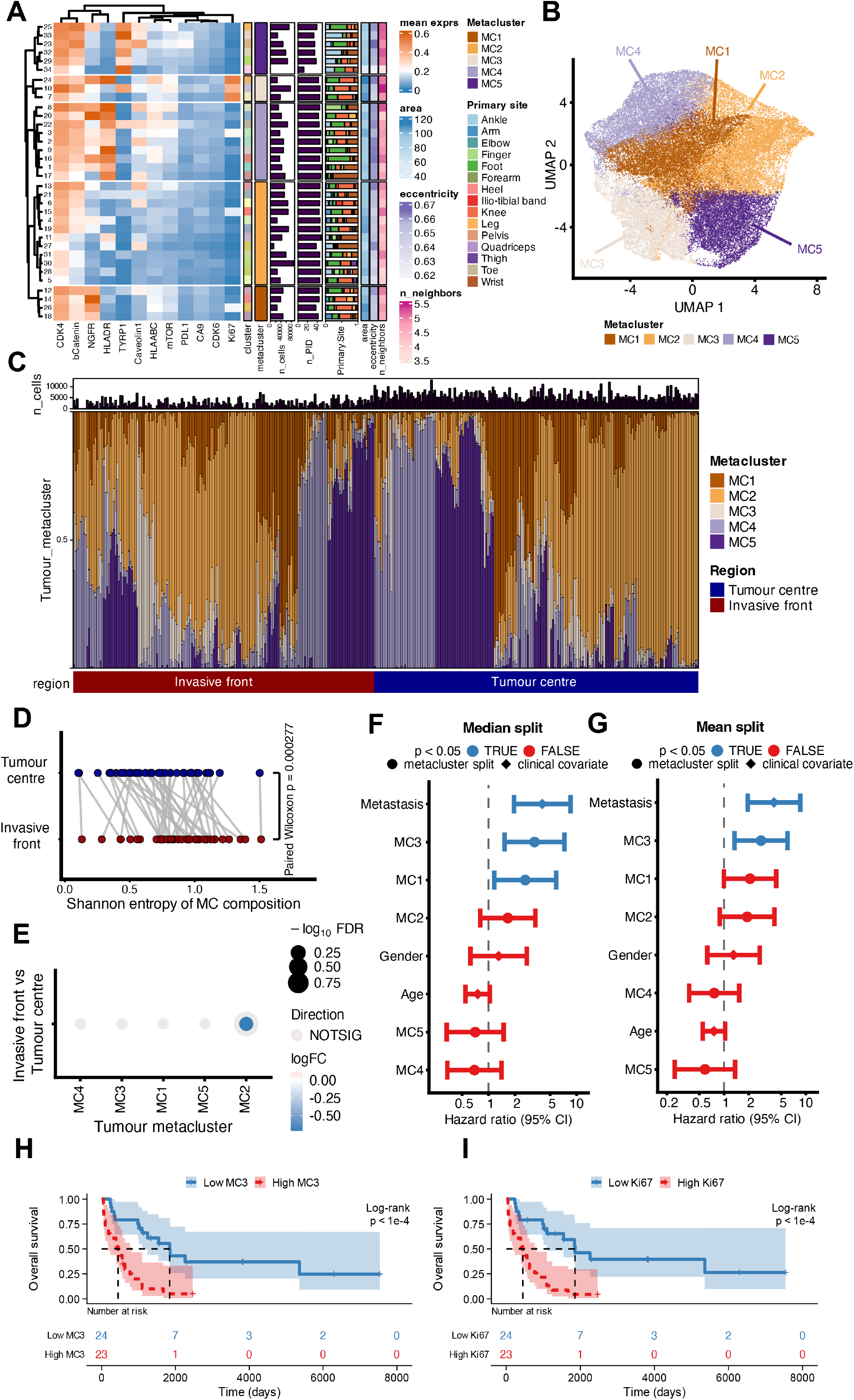
A tumour-intrinsic metacluster is associated with CCS outcome. **(A)** Heatmap of mean marker expression per tumour cluster. Rows are split by metacluster. Annotation columns to the right of the heatmap are, in order: the cluster identifier; the number of cells and the number of patients contributing to that cluster; stacked bar plots of the fraction of cells from each primary anatomic site; and the per-cluster mean of three segmentation-derived single-cell measures (cell area, eccentricity and number of neighbouring cells). The annotation columns are descriptive only: all markers shown were used for the graph-based clustering. **(B)** UMAP of 50,000 tumour cells coloured by metacluster. **(C)** Stacked bar plots of the per-ROI composition of tumour metaclusters, ordered by the proportion of metaclusters. The bar plot above shows the number of cells per ROI and the ROI type is indicated below. **(D)** Dot plot showing the Shannon entropy of tumour metaclusters per patient, comparing tumour centre and invasive front. Paired samples are joined by grey lines. P values are indicated (paired Wilcoxon signed-rank test). **(E)** Differential abundance of tumour metaclusters between invasive front and tumour centre determined by edgeR quasi-likelihood F-test. Dot size indicates −log10(FDR), colour indicates log2 fold change (invasive front vs. tumour centre), and the ring marks metaclusters called at FDR below 0.1 with Benjamini-Hochberg correction. Red, enriched at the invasive front; blue, enriched in the tumour centre; light grey, not significant. **(F and G)** Hazard ratios of overall survival from Cox proportional-hazards models adjusted for age, sex and metastatic status at diagnosis with each metacluster fraction dichotomized (F) at its median and (G) at its mean. Points indicate hazard ratio, and bars indicate 95% confidence interval. Benjamini-Hochberg correction over the five metaclusters; 47 patients with 35 events. **(H and I)** Kaplan-Meier estimates of survival by median split of (H) the MC3 fraction and (I) mean tumour-cell Ki-67 with number-at-risk tables. The indicated P values were calculated using the log-rank test; 47 patients, 35 events.

We then asked whether specific tumour MCs were associated with patient outcomes. In Cox proportional-hazards models adjusted for age, sex and metastatic status at diagnosis, with each MC fraction dichotomized at either its median or its mean, a high MC3 fraction and metastatic status were associated with shorter OS at both cut-points (**Fig. 3F–G**). Metastatic disease at diagnosis was numerically more common in MC3-high than in MC3-low patients (Fisher’s exact test, adjusted P = 0.057), but high MC3 remained associated with shorter OS after adjustment for metastatic status. Kaplan-Meier estimates confirmed the association between MC3 and OS (**Fig. 3H**). The association was also present when MC3 was kept continuous rather than dichotomized (P = 0.041; adjusted model, see Methods). Consistent with the high Ki-67 expression that characterized MC3, mean tumour-cell Ki-67 expression was also associated with OS (**Fig. 3I**). Because high Ki-67 expression defines MC3, the two analyses capture the same underlying proliferation signal, with the Ki-67 analysis expressing it in terms of a routinely measured marker (**Fig. 3I**). We found no evidence of a correlation between MC3 and any individual immune cell population (**Fig. S8E**) or of MC co-occurrence beyond that expected from compositional constraints (**Fig. S8F–G,** see Methods).

The five tumour metaclusters were also detected in the external validation cohort (**Fig. S6G–J**). In this cohort, both the proliferative MC3 metacluster and Ki-67 expression were associated with poorer overall survival (**Fig. S6M–N**); however, these associations did not reach statistical significance, likely owing to the limited number of survival events in the external cohort (10 events in 13 patients) and the resulting low statistical power.

In summary, a proliferative tumour phenotype is associated with shorter overall survival of CCS patients.

### A proliferation-associated proteomic signature is present in CCS patients with poor outcomes

Next, to extend our analysis beyond the markers measured by IMC, we used DVP, that is, LC–MS/MS proteomic profiling of laser-microdissected tissue, to characterize global proteomic signatures in CCS. For this, we used the IMC data to guide the selection of tumour regions from the same FFPE TMAs for laser microdissection and subsequent proteomic profiling, selecting a subset of patients from our cohort stratified by the MC3 fraction defined in the IMC analysis: patients were sampled from above and below the MC3 median, which in this cohort also separates shorter from longer overall survival (**Fig. 3H; Fig. S9**). An initial pilot experiment involving four patients (42 cores: 26 tumour and 16 stromal regions) confirmed the suitability of the material for untargeted mass spectrometry-based proteomics and revealed coherent, compartment-specific biological signals (**Fig. S10A–G**). In the final experiment, 7,276 proteins quantified across 141 cores from 21 patients were retained for downstream analysis (**Fig. 4A; Fig. S9**).

**FIGURE 4.**
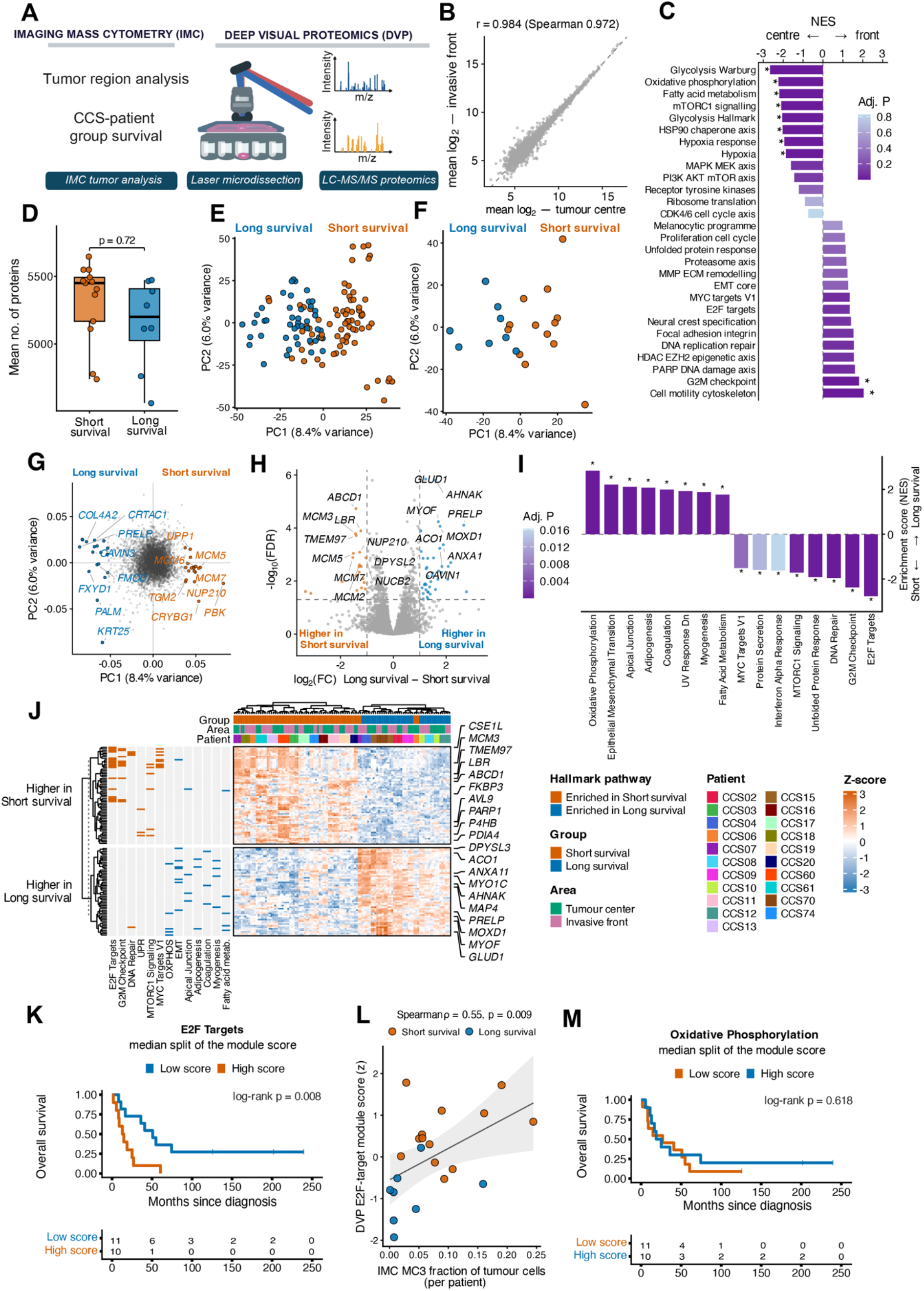
A global proteome signature characterizes CCS patients with poor outcomes. **(A)** Schematic of the deep visual proteomics (DVP) workflow: selection of tumour regions, laser microdissection, and LC–MS/MS proteomics (created with BioRender.com). **(B)** Correlation analysis of protein abundance at the invasive front vs. the tumour centre (6,616 proteins; patient-paired medians of the 21 patients with samples from both regions). Pearson r and the Spearman rank correlation coefficient rho over all proteins are indicated. **(C)** Gene-set enrichment analysis of all quantified proteins, ranked by the paired moderated t statistic for invasive front vs. tumour centre against the pathway gene sets. Bars show the normalized enrichment score, colour denotes the adjusted P value, and asterisks mark an adjusted P < 0.05. **(D)** Box plots of the mean number of proteins identified per patient by survival group. Box plots show the median and IQR, and whiskers extend to the most extreme values within 1.5×IQR from the quartiles. P values are indicated (Wilcoxon rank-sum test). **(E and F)** PCAs of the deep visual proteomics data (n = 117 cores from 21 patients), coloured by survival group. Each point represents (E) a single core or (F) the averaged core-level values for each patient (n = 21 patients). **(G)** Loadings of the individual proteins on PC1 and PC2 of the PCAs shown in panels E and F: each point is one protein, and its position shows how strongly, and in which direction, it drives each component. Top-loading proteins are labelled. **(H)** Volcano plot of differential protein expression in samples from patients with long vs. short survival (117 cores from 21 patients). Proteins called at FDR below 0.05 and an absolute log2 fold change of at least 1 are coloured and labelled. Benjamini-Hochberg correction over all tested proteins. **(I)** Gene-set enrichment analysis of all quantified proteins, ranked for long vs. short survival, against the pathway gene sets. Bars point towards the group in which each pathway is enriched. Bars show the normalized enrichment score, colour denotes the adjusted P value, and asterisks mark an adjusted P < 0.05. **(J)** Hierarchical clustering of the 120 top differential proteins between survival groups (z-scored; one column per core), annotated by survival group, region and patient (21 patients). Side bar on the left indicates the Hallmark pathway of each protein. **(K)** Kaplan-Meier curves of overall survival for patients split at the median of the E2F-targets module score with number-at-risk table. The hazard ratio was estimated using a Cox proportional hazards model. The indicated P values were calculated using the log-rank test. **(L)** Correlation analysis of the MC3 fraction of tumour cells with the E2F-target module score per patient (21 patients). The Spearman rank correlation coefficient rho and its P value are indicated; the line shows a linear fit with a 95% confidence band. **(M)** Kaplan-Meier curves of overall survival for patients split at the median of the oxidative-phosphorylation module score with number-at-risk table. The hazard ratio was estimated using a Cox proportional hazards model. The indicated P values were calculated using the log-rank test.

Paired tumour-centre and invasive-front regions from all 21 patients with both regions profiled, pooled across tumour MCs, showed similar proteomic profiles (**Fig. S10H–J**) and strong overall correlation (**Fig. 4B**). However, pathway-level analysis revealed enrichment of metabolic pathway terms in the tumour centre, including glycolysis/Warburg metabolism, oxidative phosphorylation, fatty-acid metabolism, mTORC1 signalling, and hypoxia responses, whereas the G2/M checkpoint and cell-motility pathways were enriched at the invasive front (**Fig. 4C; Fig. S10K–L**). These findings are consistent with spatially compartmentalized pathway activity in CCS tumours.

Next, to investigate associations with survival, we compared the proteomes of patients stratified by the median MC3 fraction from the IMC analysis into MC3-high/short-OS and MC3-low/long-OS groups (**Fig. 4A, Fig. S9).** Protein coverage was comparable between the two subgroups (**Fig. 4D**). Notably, the CCS proteomes separated according to these survival groups in a principal component analysis (PCA), when analysed at the level of individual TMA cores (**Fig. 4E**) and when biological replicates were aggregated per patient (**Fig. 4F**). PC1, which separated the survival groups and explained 8.4% of total variance, was primarily driven by proteins associated with cell-cycle progression, DNA replication, and proliferation (the minichromosome maintenance complex MCM2–MCM7, PBK, KPNA2, CDK1, TOP2A, and NCAPG) in the short-survival group, consistent with a proteomic proliferation signature in MC3-enriched tumours. In contrast, the long-survival group was enriched in proteins involved in mitochondrial and metabolic processes (CA5B, FABP3, MOXD1, and GLUD1) (**Fig. 4G**). Differential expression analysis revealed survival group-specific protein enrichments (**Fig. 4H**). Protein-set enrichment analysis showed that short-survival proteomes were enriched for the terms G2/M checkpoint, E2F targets, DNA repair, and unfolded protein response, whereas long-survival proteomes were enriched for oxidative phosphorylation and epithelial–mesenchymal transition (**Fig. 4I**). Unsupervised hierarchical clustering based on the most differentially abundant proteins also separated the two survival groups (**Fig. 4J**).

Next, to assess associations with survival, we calculated protein module scores for all Hallmark pathways with sufficient quantified proteins and tested each score in a separate Cox model. A higher E2F-target protein module score was associated with shorter overall survival (**Fig. 4K**). The E2F-target module score also correlated with the proportion of metacluster MC3 in the same patients (**Fig. 4L**), linking the proteomic proliferation signature to the IMC-defined MC3 state. The oxidative phosphorylation module, which had the highest enrichment score in the long-OS subgroup, was not associated with survival (**Fig. 4M**).

In summary, spatial profiling with DVP extended the characterization of the IMC-defined MC3 state from a single proliferation marker to a proteome-wide proliferation signature that includes replication licensing (MCM2–MCM7), cell-cycle regulation, DNA repair, and unfolded protein response.

### H&E-stained images of CCS tumours contain prognostic information

Because MC3 is measured by IMC, which is not available in routine diagnostics, we next asked whether the MC3 state can be recognized in routine H&E-stained slides, which would make it usable for future clinical decisions.

We trained machine-learning models to predict, from H&E images alone, whether a patient’s tumour had a low or high MC3 fraction (below or above the median used in **Fig. 3H**). For this analysis, we used H&E-stained sections consecutive to the IMC-profiled samples. Because pathology foundation models analyse small image tiles rather than whole cores, each core image was divided into non-overlapping tiles (patches), and each patch was converted into a feature vector by Phikon-v2 (Filiot et al., 2024), using the TRIDENT pipeline (Zhang et al., 2025) (**Fig. 5A**). The patch features of each core were then combined and used to classify the core as MC3-low or MC3-high with three machine-learning approaches: support vector machine (SVM), random forest, and an attention-based multiple-instance learning (MIL) model (Attn-MIL) (**Fig. 5A**). Before classification, we removed patches without tumour cells. The remaining patch feature vectors were grouped into ten clusters of similar morphology by k-means clustering (**Fig. S11A–D**). Representative patches of each cluster were manually inspected, and the one cluster consisting predominantly of non-tumour tissue was excluded (**Fig. S11A**).

**FIGURE 5.**
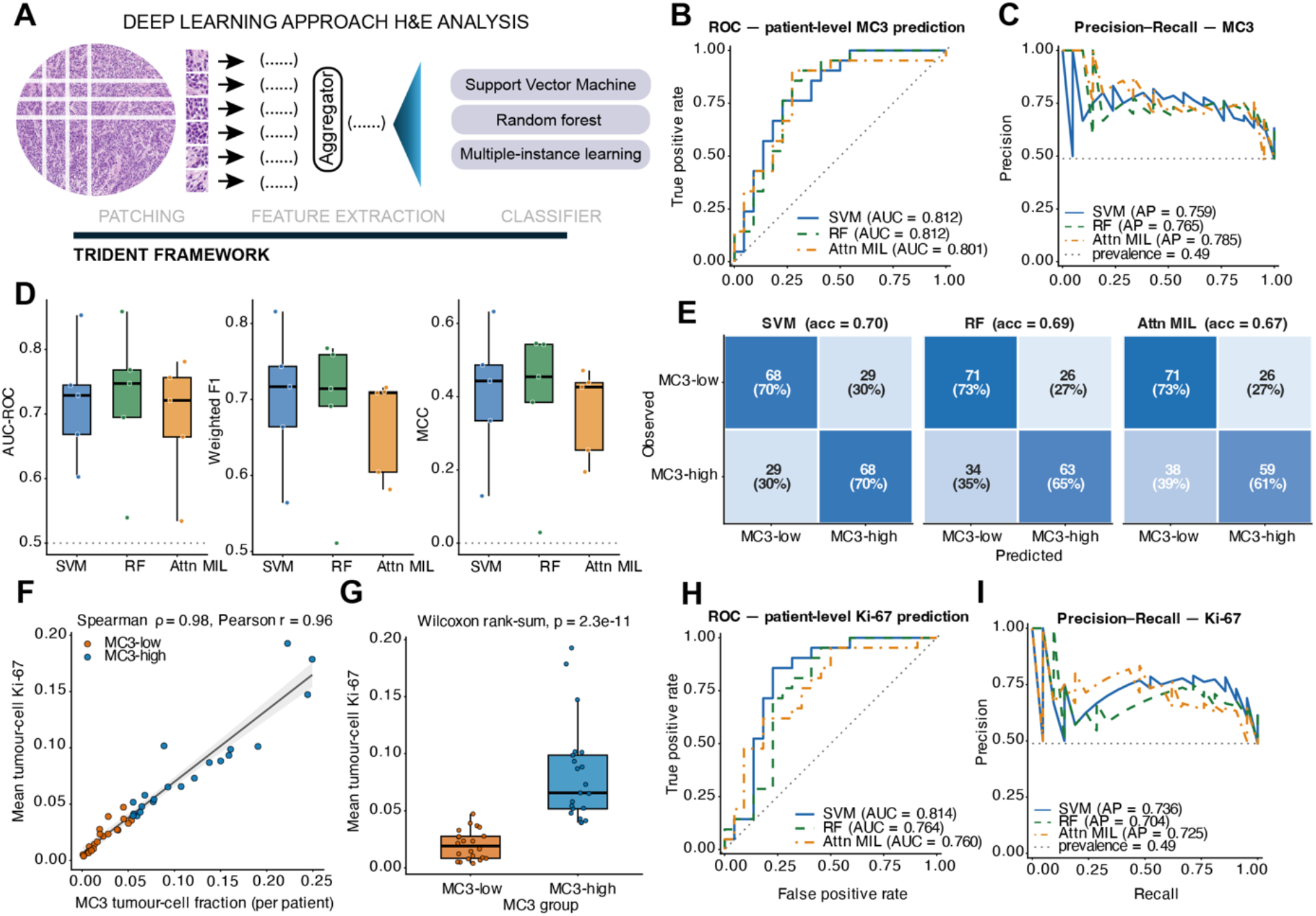
H&E morphology predicts the proliferative tumour metacluster in CCS. **(A)** Schematic of the analysis of H&E-stained tumour sections within the TRIDENT framework. H&E images were divided into non-overlapping patches; each patch was converted into a feature vector by the pathology foundation model Phikon-v2. The patch features of each core were combined (mean and maximum for SVM and RF; attention-weighted for Attn-MIL) and used for classification with either a support vector machine (SVM), a random forest (RF) or an attention-based multiple-instance learning (MIL) model (Attn-MIL). **(B)** ROC curves for the prediction of patient-level low vs. high tumour MC3 abundance. Curves were computed on out-of-fold predictions from stratified five-fold cross-validation grouped by patient; the area under the curve is indicated for SVM, random forest (RF), and Attn-MIL models. **(C)** Precision-recall curves for the same task. The average precision is indicated per model, and the positive-class prevalence of 0.49 at patient level is shown as the dotted baseline. **(D)** Box plots of fold-wise performance of each model across the five patient-grouped cross-validation folds, reported as AUC-ROC, weighted F1 score, and Matthews correlation coefficient (MCC). Box plots show the median and IQR, and whiskers extend to the most extreme values within 1.5×IQR from the quartiles. Points denote individual folds. **(E)** Confusion matrices from out-of-fold predictions at a probability threshold of 0.5, calculated per core. Rows are observed class; columns are predicted class. Each cell gives the number of cores and that number as a percentage of all cores of the same observed class (each row sums to 100%), and the core-level accuracy is indicated per model. **(F)** Scatter plot comparing the mean tumour-cell Ki-67 expression with the MC3 tumour-cell fraction, per patient, coloured by MC3 group. Spearman rho and Pearson r are indicated; linear smoothing with a 95% confidence interval is displayed in grey. **(G)** Box plots of the mean tumour-cell Ki-67 expression by MC3 group. Box plots show the median and IQR, and whiskers extend to the most extreme values within 1.5×IQR from the quartiles. Points denote individual patients. P values are indicated (Wilcoxon rank-sum test). **(H, I)** As in B and C, for the prediction of high vs. low mean tumour-cell Ki-67 expression, dichotomized at the cohort median.

All three models predicted low versus high tumour MC3 abundance above chance (patient-level area under the receiver operating characteristic curve (AUC-ROC) 0.81 for SVM, 0.81 for random forest, and 0.80 for Attn-MIL; 95% bootstrap confidence intervals 0.67–0.93, 0.66–0.94 and 0.64–0.93, respectively), and all three exceeded a null distribution obtained by permuting the patient labels 2,000 times (**Fig. 5B–C**). Performance varied across folds, especially for Attn-MIL (fold-wise AUC-ROC 0.60–0.85 for SVM, 0.54–0.86 for random forest, and 0.53–0.78 for Attn-MIL), consistent with the modest cohort size (**Fig. 5D**). At a 0.5 threshold, all models classified MC3-low versus MC3-high H&E cores with distinct sensitivity profiles (**Fig. 5E**).

Because MC3 is associated with high Ki-67 expression, we next asked whether its abundance could be read directly from this marker, which is routinely quantified in clinical practice. As expected, per-patient mean tumour-cell Ki-67 expression correlated closely with MC3 abundance (**Fig. 5F–G**). Because Ki-67 is in effect the defining marker of MC3, the higher Ki-67 in MC3-high patients follows from how the groups are defined rather than being an independent finding. H&E classifiers trained on the Ki-67 median split therefore performed similarly to those trained on the MC3 split (patient-level AUC-ROC 0.81, 0.76, and 0.76 for SVM, random forest, and Attn-MIL, respectively; **Fig. 5H–I**), indicating that the morphological signal is linked to Ki-67 expression.

Together, these results show that H&E morphology identifies the IMC-defined, poor-prognosis tumours with Ki-67 expression serving as a potential surrogate marker.

### Tumour composition and proteome are largely preserved between primary tumours and lymph-node metastases

To assess whether primary and metastatic tumours resemble one another, we compared tumour metacluster composition and proteome profiles between matched primary tumours and lymph-node metastases in paired samples from our cohort (n = 11 patients).

Primary tumour phenotypic compositions were broadly conserved at the metastatic sites. The pairwise correlation of tumour metaclusters was similar within primary tumours, between primary and metastatic samples, and within metastases (**Fig. 6A**). Further, PCA on the tumour metacluster composition showed substantial overlap between primary and metastatic samples (**Fig. 6B**). Compositional diversity was also similar between sites, with no differences in any of the indices examined (**Fig. 6C**). The proliferative metacluster MC3 was more abundant in metastases in 8 of 11 patients, although this was not statistically significant (**Fig. 6D–E).** Similarly, proteomic profiles of paired samples showed no clear separation by PCA (n = 9 patients) (**Fig. 6F**; **Fig. S10M–O**). However, we identified 42 proteins as differentially abundant between primary and metastatic samples (**Fig. 6G**); the protein most significantly enriched in metastasis was SERPINB9 (**Fig. 6G, H**), an intracellular serine protease inhibitor previously described as promoting metastasis in different types of neoplasm (Bian et al., 2024; Huang et al., 2024). A separate paired analysis yielded a similar result, with higher SERPINB9 abundance in metastases than in matched primary tumours in 8 of 9 patients. Further, gene-set enrichment analysis revealed enrichments of proliferation- and cell-cycle-related programmes, the proteasome axis, and the G2/M checkpoint pathway in metastases and of the oxidative phosphorylation pathway in primary tumours (**Fig. S10P).**

**FIGURE 6.**
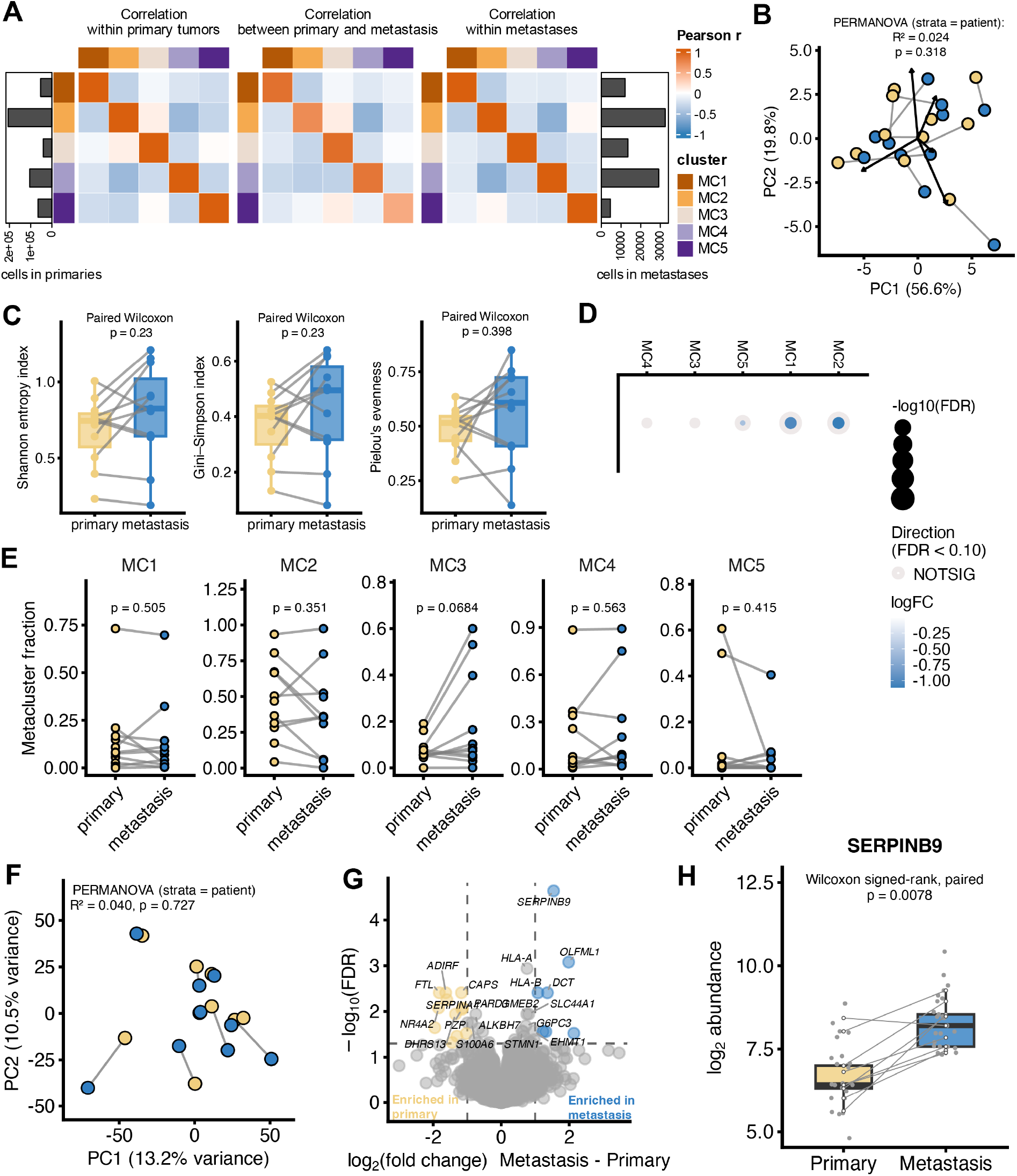
Tumour metacluster composition and global proteome are largely preserved between primary CCS tumours and lymph-node metastases. **(A)** Pearson correlation matrices of tumour metacluster fractions (MC1–MC5), calculated within primary tumours, between matched primary tumours and metastases, and within metastases (11 patients with matched samples). Side bar plots indicate the number of cells assigned to each metacluster in primary tumours and in metastases. **(B)** PCA of tumour metacluster composition, showing primary tumours and metastases (11 patients); matched samples are connected by grey lines. The PERMANOVA result, stratified by patient, is indicated. **(C)** Box plots of the diversity of tumour metacluster composition in matched primary and metastatic samples, assessed using the Shannon entropy index, the Gini-Simpson index, and Pielou’s evenness (11 patients). Box plots show the median and IQR, and whiskers extend to the most extreme values within 1.5×IQR from the quartiles. Points show individual samples, and lines connect matched pairs. P values are indicated (paired Wilcoxon signed-rank test). **(D)** Differential abundances of tumour metaclusters between metastasis vs. primary tumour determined by edgeR quasi-likelihood F-test (11 patients). Dot size indicates −log10(FDR), colour indicates log2 fold change (metastasis vs. primary tumour), and the ring marks metaclusters called at FDR below 0.1 with Benjamini-Hochberg correction. **(E)** Dot plots showing paired comparisons of metacluster fractions in matched primary and metastatic samples (11 patients). Lines connect matched samples. Unadjusted P values are indicated (paired Wilcoxon signed-rank test). **(F)** PCA of proteomic data from primary tumour vs. metastasis samples (9 patients); matched samples are connected by grey lines. The PERMANOVA result, stratified by patient, is indicated. **(G)** Volcano plot of differential protein abundance in metastasis vs. primary tumour (47 cores from 9 patients). Proteins called at FDR below 0.05 and an absolute log2 fold change of at least 1 are coloured and labelled. Benjamini-Hochberg correction over all tested proteins. **(H)** Box plot of SERPINB9 abundance in matched primary tumours and metastases. Grey points show the cores; white points show the identifier-level median per patient connected by lines. Box plots show the median and IQR, and whiskers extend to the most extreme values within 1.5×IQR from the quartiles. P values are indicated (paired Wilcoxon signed-rank test).

IMC-based tumour metacluster composition was largely preserved between primary tumours and lymph-node metastases. At the proteome level, a small number of proteins, including SERPINB9, showed differential abundance between these sample types, and metastatic samples showed a trend towards proliferative and proteasome-related programmes.

### c-MET and B7-H3 are potential targets of ADCs in CCS

Therapeutic options for advanced-stage CCS remain limited. We therefore investigated whether CCS tumour cells express surface antigens that could serve as targets for ADCs.

Our IMC panels included 15 antigens that have previously been evaluated in clinical trials and preclinical models of solid tumours (HER2, HER3, c-MET, FOLR1, MUC16, cadherin-6, LY6E, TROP2, EGFR, NaPi2b, tissue factor, claudin-6, AXL, CD70 in panel II; B7-H3 in panel I; Fig. S3H–M). Across individual cores, there was heterogeneous expression of these proteins on tumour cells with B7-H3 and c-MET exhibiting the highest expression levels (**Fig. 7A**). To rank the target antigens, we scored each on five criteria: tumour selectivity, expression homogeneity, patient coverage, percentage of tumour cells positive for the antigen, and median expression on tumour cells. Each score was min-max normalized to 0–1 across all 15 antigens, and the five scaled values were averaged into a composite score. This analysis identified B7-H3 (composite score 0.91) and c-MET (composite score 0.68) as the two highest-ranking targets (**Fig. 7B**); expression of both was then confirmed visually (**Fig. 7C**). For both proteins, tumour-cell expression did not differ significantly between the tumour centre and the invasive front and was correlated between the two regions (**Fig. 7D–E**), indicative of spatially stable target availability within tumours. In the matched primary and metastatic samples, expression of neither antigen differed significantly (**Fig. 7F**). Both antigens were also detectable by immunohistochemistry (**Fig. 7G**). Finally, neither B7-H3 nor c-MET was associated with overall survival (**Fig. 7H–I**), an important consideration when evaluating clinical relevance as the target of an ADC.

**FIGURE 7.**
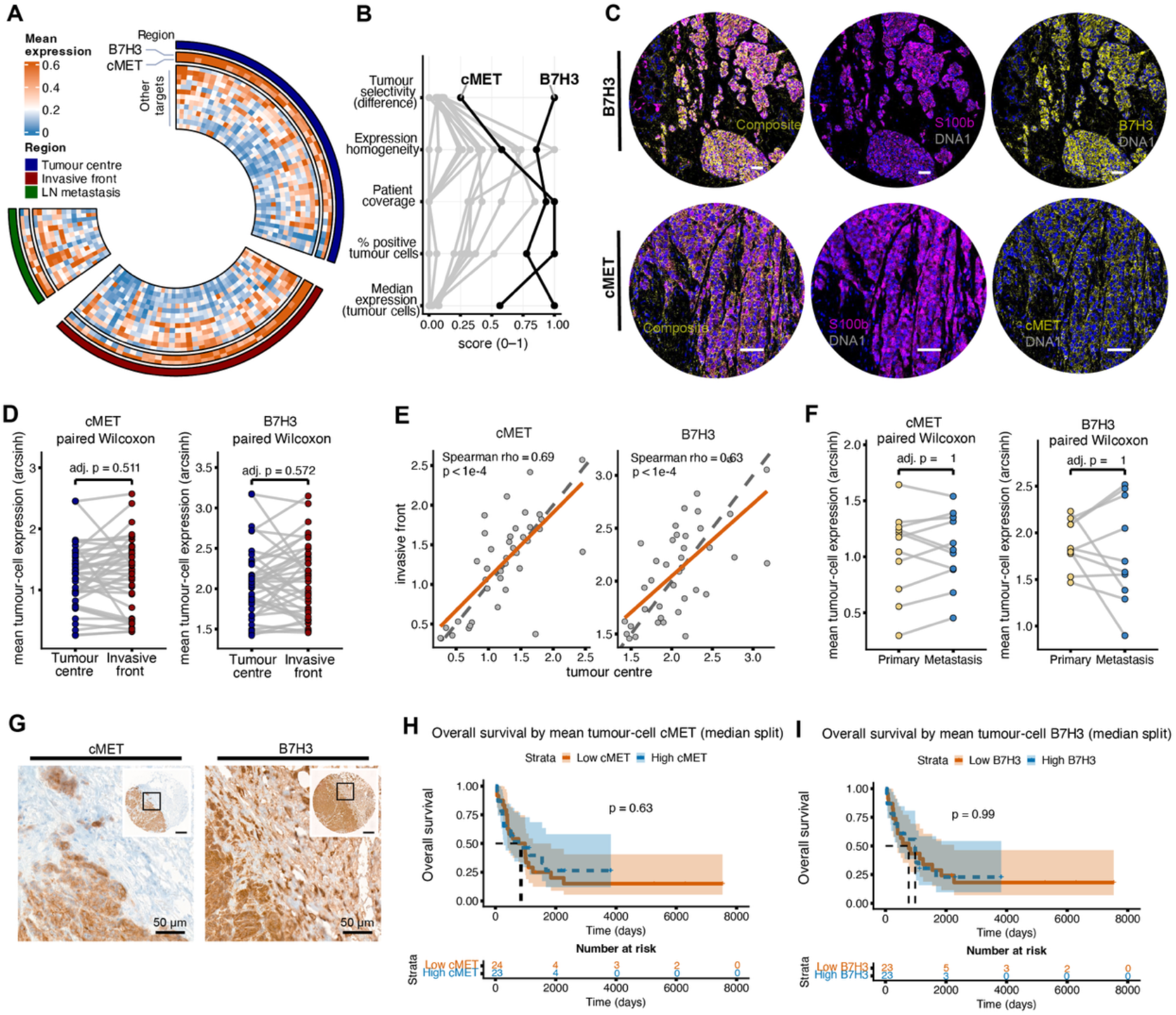
c-MET and B7-H3 as therapeutic candidates in CCS. **(A)** Circular heatmap with one slice per region per patient (tumour centre, invasive front, lymph-node metastasis). Rings show the mean tumour-cell expression of the 15 ADC target antigens, rescaled within each ring. c-MET and B7-H3 are highlighted. **(B)** Composite target ranking of the 15 antigens according to the indicated five criteria, each min-max scaled to 0–1 across the antigens. **(C)** Representative ROIs for B7-H3 (top) and c-MET (bottom) expression, shown as a composite image (left), as S100b with DNA (middle), and as the candidate marker with DNA (right). Scale bars, 50 µm. **(D)** Dot plots comparing mean tumour-cell expression of c-MET and B7-H3 between paired tumour centre and invasive front samples, paired per patient. Lines connect the two regions of a patient. P values are indicated (paired Wilcoxon signed-rank test). **(E)** Correlation analysis of the mean tumour-cell expression of c-MET and B7-H3 between the tumour centre and invasive front. The Spearman rank correlation coefficient rho and its P value are indicated; the dashed line shows the identity and the orange line a linear fit. **(F)** Dot plots comparing the mean tumour-cell expression of c-MET and B7-H3 between matched primary tumours and metastases. Lines connect the two sample types per patient. P values are indicated (paired Wilcoxon signed-rank test). **(G)** Representative immunohistochemistry for c-MET and B7-H3. Scale bars, 50 µm. **(H, I)** Kaplan-Meier curves of overall survival for patients stratified based on tumour-cell expression of (H) c-MET and (I) B7-H3 with number-at-risk tables. The indicated P values were calculated using the log-rank test; 47 patients, 35 events. Patients were stratified by a median split of the mean marker expression per tumour cell.

These data nominate c-MET and B7-H3 as candidate ADC targets in CCS.

## DISCUSSION

CCS is a rare and aggressive soft-tissue sarcoma (Cornillie et al., 2016; Ikuta et al., 2023; Wetterwald et al., 2023). To characterize the microenvironment of this type of tumour, we analysed samples from a multicentre cohort of 52 CCS patients resulting in a single-cell, spatially resolved proteomic atlas of the disease. Together with global proteomic analysis of laser-captured tissue, this atlas enabled detailed spatial characterization of the tumour and immune compartments in CCS and assessment of their associations with clinical outcome.

We showed that CCS tumours are immune-cold with sparse, myeloid-dominated immune infiltrates especially in the tumour centre, where macrophages are the main immune cells interacting with tumour cells. Consistent with the immune-low/immune-desert sarcoma classes described by Petitprez et al. (Petitprez et al., 2020), which respond poorly to checkpoint blockade, we observed few lymphoid features and no tertiary lymphoid structures in tumours from this CCS cohort.

In contrast to studies on carcinoma, where a myeloid-to-lymphoid gradient with lymphoid enrichment at the tumour margin has been reported in colorectal cancer (Lin et al., 2023) and hepatocellular carcinoma (Ruf et al., 2023), the immune organization at the invasive front remains virtually unexplored in sarcomas. We made use of our unique dataset to study this. Our analysis revealed spatial compartmentalization of the immune infiltrate in CCS. The invasive front was consistently more immune-diverse than the tumour centre, with higher immune entropy, greater lymphoid density, and a broader spectrum of T-cell states, including exhausted PD-1⁺ and TCF1/7⁺ naïve-like T cells. In contrast, the tumour centre was dominated by myeloid cells, mainly CD68⁺ macrophages and CD163⁺ HLA-DRhigh or HLA-DRlow macrophages; together, these accounted for about three-quarters of all tumour–immune cell contacts in these tumours. Notably, once the regional difference in abundance was accounted for, the infiltration score of all three macrophage populations was comparable between the tumour centre and the invasive front, indicating that the higher relative myeloid presence in the tumour centre is carried by macrophage abundance rather than by a difference in how closely macrophages engage tumour cells. Overall, the spatial compartmentalization of immune cells suggests that there is some feature of CCS tumours that acts as a barrier to immune infiltration, with myeloid-lineage cells being the main immune population able to enter the tumour core. However, whether this reflects a physical barrier formed by tumour cells or by stroma, chemokine gradients, or vascular access cannot be resolved from these data. To our knowledge, this is the first study to characterize the immune-cell composition at the invasive front in a sarcoma subtype.

Our IMC analysis further identified distinct tumour cell phenotypes within CCS, including a proliferative cell type associated with poor outcome. In contrast, immune composition, whether at the invasive front or in the tumour centre, showed no robust association with outcome. Our DVP analysis extended the characterization of the IMC-defined MC3 proliferative tumour cell state from a single marker to a broader proteome-wide signature involving replication licensing (MCM2–MCM7), cell-cycle regulation, DNA repair, and the unfolded protein response. Together, these complementary modalities support the presence of a subgroup of CCS patients characterized by high proliferative activity and poor clinical outcome. As no established classification or grading system exists for CCS, Ki-67 may serve as a simple, clinically applicable surrogate marker for the proliferative cell type and its associated proteome-wide proliferation signature. Furthermore, we showed that this prognostic information was also partially captured by standard H&E images, further paving the way for personalized patient stratification.

From a therapeutic perspective, our atlas indicates that CCS lacks key features typically associated with checkpoint inhibitor response: T cells are sparse, and macrophages dominate the immune compartment. This is consistent with the limited activity of immune-checkpoint blockade in CCS, including in the OSCAR trial (NCCH1510; trial registration UMIN000023665), in which nivolumab achieved no objective responses among the 12 CCS patients enrolled (Nishikawa et al., 2024). Our data are also consistent with the limited activity of PD-1/PD-L1 inhibitors across sarcoma (D’Angelo et al., 2018; Tawbi et al., 2017). Given the myeloid-rich immune microenvironment observed in our cohort, strategies that target macrophage-mediated immune exclusion, such as CSF1R inhibition combined with PD-1 blockade (Zhu et al., 2014), warrant investigation. We note that very recently, the single-arm phase II IMMUNOSARC II trial (NCT03277924) demonstrated clinical activity of sunitinib combined with nivolumab in advanced CCS with partial responses in 3 of 21 radiologically evaluable patients (14.3%), a median progression-free survival of 6.2 months, and a 6-month progression-free survival rate of 50.1% (Martin-Broto et al., 2026). These findings support further investigation of antiangiogenic therapy combined with checkpoint blockade; however, the non-randomized, single-arm design precludes determining the contribution of either agent to the observed activity. In our cohort, the presence of PD-1⁺ and TCF1/7⁺ T cell populations at the invasive front may offer a biological rationale for response in a subset of CCS patients. Because none of the cases included in this study received such drugs, treatment response could not be assessed. Antigen-directed therapies such as bispecific T cell engagers have recently emerged as potential treatment strategies for CCS. Unlike checkpoint inhibitors, T cell–redirecting therapies can target tumour-restricted antigens. Based on this rationale, tebentafusp, a gp100–HLA-A*02:01-directed CD3-directed T-cell engager with clinical activity in metastatic uveal melanoma (Nathan et al., 2021), is being evaluated in a phase II trial (NCT06942442) in patients with HLA-A*02:01-positive unresectable or metastatic CCS.

Our data nominate c-MET and B7-H3 as candidate antigen-directed therapeutic targets in CCS. Because antibody–drug conjugate activity depends primarily on cell-surface antigen expression rather than inhibition of the target’s signaling function, ADCs may be effective even when the antigen is not a key oncogenic driver (Drago et al., 2021; Tarantino et al., 2022). They may also retain activity at antigen-expression levels below the thresholds used for kinase-directed therapies (Modi et al., 2022). Relevant to this, trastuzumab deruxtecan, an ADC targeting HER2, has shown activity in desmoplastic small round cell tumour (Slotkin et al., 2026), which, like CCS, is a fusion-driven rare sarcoma. c-MET has been shown to be upregulated in CCS (Davis et al., 2010; Ikuta et al., 2023). The prospective EORTC 90101 ‘CREATE’ trial (NCT01524926) evaluated the MET inhibitor crizotinib in advanced CCS and reported limited clinical activity, with few objective responses by RECIST (Schöffski et al., 2017). Given the limited clinical activity of crizotinib in CCS, an alternative strategy would be to use an ADC that binds cell-surface c-MET to deliver a cytotoxic payload. Telisotuzumab vedotin, a c-MET-directed ADC, has shown efficacy in different types of tumours (Camidge et al., 2024; Wang et al., 2017). In CCS, we observed uniformly high c-MET expression across the tumour centre and invasive front and in both primary and metastatic samples. This consistency supports investigating c-MET as a target for ADC-based approaches. Our other proposed target, B7-H3, is broadly expressed across soft-tissue sarcomas (Lynch et al., 2024), and B7-H3-directed agents are in clinical development: the antibody–drug conjugate ifinatamab deruxtecan achieved a confirmed objective response rate of 48.2% in previously treated extensive-stage small-cell lung cancer (Rudin et al., 2026). CCS is listed among the eligible diseases of an ongoing paediatric and young-adult phase I B7-H3 CAR T-cell trial (NCT04483778). As for c-MET, our study provides the spatial expression pattern of this target, demonstrating homogeneous expression across tumour regions and between primary and metastatic samples. Also, neither B7-H3 nor c-MET expression was associated with overall survival, so selecting patients on target expression would not by itself enrich for a prognostic subgroup, a point relevant to the design and interpretation of potential clinical trials.

In conclusion, our single-cell spatial proteomic analysis identifies CCS as an immune-cold, myeloid-dominated tumour with a spatially compartmentalized immune infiltrate and a Ki-67-high tumour cell state associated with both a related proteomic signature and poorer survival. Further, we propose c-MET and B7-H3 as candidate targets for antibody–drug conjugates and other antigen-directed approaches in CCS.

This study has limitations. First, the number of patient samples was limited due to the inherent challenges of studying ultra-rare neoplasms like CCS, for which high-quality, clinically annotated tissue is extremely difficult to obtain. Therefore, this study should be interpreted in the context of its exploratory design. Second, the spatial analyses relied on TMA cores, which sample only a limited fraction of the tumour and may therefore underestimate intratumoural heterogeneity. To mitigate this limitation, multiple cores were analysed for all cases. Third, as this study was based on retrospectively collected human samples, it was descriptive in nature and could identify associations but not establish causality. Fourth, the prognostic associations of MC3 abundance and tumour-cell Ki-67 expression require independent validation before clinical application. The median cut-offs used to define high and low groups were exploratory and derived from the study cohort. Future studies should use a prespecified scoring protocol to assess tumour-cell Ki-67 by immunohistochemistry and quantify mitoses on whole-slide sections in a large, independent CCS cohort. Finally, and more broadly, these limitations reflect structural challenges inherent to research in ultra-rare sarcomas rather than weaknesses specific to the present study. Patient-partnered initiatives have demonstrated the potential to accelerate recruitment in this setting (Painter et al., 2020). Looking forward, prospective, standardized collection of CCS tissue alongside plasma and blood samples will be an important next step towards a more comprehensive understanding of CCS. The present study, conducted within the Clear Cell Sarcoma Consortium initiative, lays the groundwork for these efforts.

## Data Availability

IMC data, proteomics data, and H&E images will be made available on zenodo as of the date of publication (10.5281/zenodo.24281886). All original code has been deposited on GitHub and is publicly available as of the date of publication (https://github.com/BodenmillerGroup/CCS_project). Any additional information required to reanalyze the data reported in this paper is available from the lead contact upon request.

## RESOURCE AVAILABILITY

### Lead contact

Requests for further information and resources should be directed to and will be fulfilled by the lead contacts, Bernd Bodenmiller and Nischalan Pillay.

### Materials availability

This study did not generate new unique reagents.

### Data and code availability

IMC data, proteomics data, and H&E images will be made available on Zenodo as of the date of publication (10.5281/zenodo.24281886). All original code has been deposited on GitHub and is publicly available as of the date of publication (https://github.com/BodenmillerGroup/CCS_project).

## ETHICS

For samples from the UK cohort: The Royal National Orthopaedic Hospital (RNOH) Biobank is approved by the National Research Ethics Committee of the Health Research Committee (reference: Integrated Research Application System (IRAS) project identifier: 272816). This study was approved by the UCL/UCLH Biobank for Studying Health and Disease – EC39.25 The Multi-Omic Diversity of Clear Cell Sarcoma. Written informed consent was obtained from all individual participants prior to inclusion in the study. For samples from the Swiss cohorts: the study was approved at the University Hospital Basel and the University Hospital Zürich following approval by the Ethikkommission beider Basel (ref. 274/12).

## ACKNOWLEDGMENTS

We thank all patients who donated tumour samples. This study was funded by the Edward Showler Foundation (https://www.edwardshowlerfoundation.com), Transcend Sarcoma (https://www.transcendsarcoma.com) and Sarcoma UK (SUK 09.2022). The Clear Cell Sarcoma Consortium originated within the FORTRESS network (https://www.kuleuven.be/fortress). We are grateful to the Bone Cancer Research Trust that provides infrastructure support for the RNOH biobank. B.B. was supported by a Swiss National Science Foundation (SNSF) project grant, an SNSF R’Equip grant, and the European Research Council (ERC) under the European Union’s Horizon 2020 research and innovation programme (grant agreement no. 866074, “Precision Motifs”). N.P. has received funding from Cancer Research UK (Award number 18387) and holds a Cancer Research UK Career Establishment award (RCCCEANov23/100003). N.D. received funding from the Nuovo-Soldati Foundation.

## AUTHOR CONTRIBUTIONS

Conceptualization, S.H., N.P., and B.B.; methodology, S.H., N.D., B.E., L.M., M.D., C.D., A.W., P.S., N.d.S., D.B., C.P., F.M., S.C., and N.P.; formal analysis, S.H., N.D., and B.E.; investigation, S.H., L.M., F.M., S.C., and N.P.; resources, B.B.; visualization, S.H.; writing – original draft, S.H.; writing – review & editing, S.H., N.D., B.E., L.M., M.D., A.F., A.I., C.D., A.W., P.S., N.d.S., D.B., C.P., F.M., S.C., N.P., and B.B.; supervision, B.B. and N.P.; funding acquisition, N.P. and B.B. S.H., N.P., and B.B. had full access to all the data in the study and take responsibility for the integrity of the data and the accuracy of the data analysis.

## DECLARATION OF GENERATIVE AI AND AI-ASSISTED TECHNOLOGIES

During the preparation of this work, the authors used Claude (Anthropic) to copyedit the manuscript. After using this tool, the authors reviewed and edited the content and take full responsibility for the content of the publication.

## SUPPLEMENTAL INFORMATION

• Figures S1–S11

• Tables S1–S7

## MATERIALS AND METHODS

### PATIENT COHORTS

#### Discovery cohort

Archival formalin-fixed, paraffin-embedded (FFPE) tumour samples and clinical data were obtained from the Royal National Orthopaedic Hospital biobank (pathology archives and London Sarcoma Service databases), the pathology service of the University Hospital Zürich, and the pathology service of the University Hospital Basel. For samples from the UK cohort: The Royal National Orthopaedic Hospital (RNOH) Biobank is approved by the National Research Ethics Committee of the Health Research Committee (reference: Integrated Research Application System (IRAS) project identifier: 272816). This study was approved by the UCL/UCLH Biobank for Studying Health and Disease – EC39.25 The Multi-Omic Diversity of Clear Cell Sarcoma. Written informed consent was obtained from all individual participants prior to inclusion in the study. For samples from the Swiss cohorts: the study was approved at the University Hospital Basel and the University Hospital Zürich following approval by the Ethikkommission beider Basel (ref. 274/12). Pathology archives were searched for sarcomas coded as “clear cell sarcoma,” “melanoma of soft tissue,” or “melanoma of soft parts.” Fifty-six cases with adequate FFPE material were identified (44 from the Royal National Orthopaedic Hospital, 8 from Zürich, and 4 from Basel). All cases were reviewed by pathologists (S.H., D.B., C.P., and N.P.), and only cases with a confirmed *EWSR1* rearrangement were included. All samples were obtained before treatment. After IMC quality control, 52 patients remained (40 from the Royal National Orthopaedic Hospital, 8 from Zürich, and 4 from Basel). Of the 52 analysed patients, 32 were recorded as female and 20 as male; median age at diagnosis was 31 years (range, 10–69) (**Sup. Table S1**). Ancestry, race, ethnicity, and socioeconomic status were not available in the archival records. No statistical method was used to predetermine sample size; all eligible cases with adequate tissue were included.

#### External validation cohort

FFPE samples from an external CCS cohort were obtained from the Laboratory of Experimental Oncology, Department of Oncology, KU Leuven, under a material transfer agreement (UZ-26/559). After quality control and exclusion of control cores containing normal tissue, 56 images from 20 patients were retained (12 male, 7 female, and 1 unrecorded; median age 40 years, range 11–71) (**Sup. Table S4**). Survival analysis included the 13 patients with usable follow-up (10 events). Ethical clearance for the use of these samples was obtained, and the local ethics requirements were followed.

#### Cutaneous melanoma cohort

A cutaneous melanoma cohort comprising 38 patients (25 male and 13 female) was profiled for comparison; these are the cases of a proprietary cohort from Navignostics (four regions of interest per patient, 152 in total). Age at sampling was available for 29 patients (median, 70 years; range, 35–95). Melanoma stage was recorded for 37 patients, ranging from IB to IV, with 19 classified as stage IIIC. TNM classification was unavailable (**Sup. Table S5**), and treatment before sampling was not recorded for any patient. Ethical clearance for the use of these samples was obtained following the local ethics requirements.

### EXPERIMENTAL PROCEDURES

#### Region selection and TMA construction

Regions of interest (ROIs) were selected by a board-certified pathologist (S.H.) and classified as invasive front, tumour centre, or lymph-node metastasis. The invasive front was defined from the macroscopic architecture of the surgical specimen, using blocks designated as tumour margin and confirmed to contain less than 50% tumour cells on visual inspection. The tumour centre was defined as a region composed predominantly of tumour cells, from blocks designated as the centre of the tumour. Three TMAs (ZTMA_297, ZTMA_298, and ZTMA_299) with 1-mm cores were constructed. Paired tumour-centre and invasive-front samples were available for 40 patients and matched primary tumour and lymph-node metastasis samples for 11 patients.

### IMAGING MASS CYTOMETRY

#### Antibody panels

Two panels were used (**Sup. Tables S2** and **S3**). Panel I characterized the immune and tumour landscape. Tumour cells were characterized with the melanocytic markers S100b, MelanA, NGFR, and TYRP1 together with markers of signalling, cell cycle, hypoxia, apoptosis, and immune modulation (β-catenin, mTOR, CDK4, CDK6, caspase-3, CA9, Ki-67, and B7-H3). Myeloid cells were characterized with CD11c, CD68, CD163, CD15, arginase-1, PD-L1, and IDO1, and lymphoid cells with CD45, CD3e, CD8a, CD4, CTLA-4, LAG-3, 4-1BB, PD-1, TCF1/7, FOXP3, CD20, CD38, CD56, and granzyme B. Tissue architecture was assessed with SMA, CD31, PDGFRβ, podoplanin, vimentin, pan-keratin, and caveolin-1, and antigen presentation with HLA-ABC and HLA-DR. Histone H3 and iridium intercalator marked nuclei. Panel II profiled ADC target antigens together with tumour, stromal, and immune markers. Candidate ADC targets were selected on the basis of antibody compatibility with IMC and of prior evaluation as ADC targets in clinical trials (Dumontet et al., 2023; Tsuchikama et al., 2024): HER2 (Modi et al., 2022), HER3 (Larsen et al., 2023), c-MET (Camidge et al., 2024), FOLR1 (Krivak et al., 2025), MUC16 (Liu et al., 2016), cadherin-6 (Patel et al., 2025), LY6E (Krah et al., 2026), TROP2 (Mullard, 2025), EGFR (Deng et al., 2026), NaPi2b (Vogl et al., 2025), tissue factor (Ren et al., 2025), claudin-6 (Katoh et al., 2025), AXL (Rohrberg et al., 2025), and CD70 (Pal et al., 2019). B7-H3 (Rudin et al., 2026) was measured in panel I. Panel II further contained SMA, pan-keratin, vimentin, CD31, S100b, MelanA, Ki-67, B2M, NGFR, TYRP1, HLA-ABC, HLA-DR, PRAME, caveolin-1, caspase-3, CA9, CD68, MPO, CD15, granzyme B, arginase-1, IDO1, PD-L1, CD45, CD20, CD3e, CD4, and CD38. **Sup. Tables S2** and **S3** list the target, metal isotope, clone, host species, clonality, isotype, supplier, catalogue and lot numbers, working concentration, conjugation method, positive and negative control samples, and titration quality-control result for each antibody conjugate.

#### Antibody conjugation and staining

Antibodies were conjugated to lanthanide metals with the Maxpar antibody labelling kit (Standard BioTools) according to the manufacturer’s instructions, quantified with a NanoDrop One spectrophotometer (Thermo Fisher Scientific), and stored at 4 °C in antibody stabilizer (Candor Bioscience). Conjugates were titrated on reference cell lines before panel assembly. TMA sections (2 μm) were deparaffinized in xylene (three times for 10 min) and rehydrated through a graded ethanol series (2 × 100% for 5 min; 2 × 96%, 90%, 80%, and 70% for 3 min each). Heat-induced antigen retrieval was performed in Tris-EDTA buffer (pH 9) at 95 °C for 30 min in a decloaking chamber (Biocare Medical). After cooling for 20 min, sections were blocked with 3% BSA and 0.1% Tween in Tris-buffered saline (TBS; 150 mM NaCl, 50 mM Tris-HCl, pH 7.6) for 1 h. To amplify the CD4 signal, sections were incubated with unlabelled rabbit anti-CD4 overnight at 4 °C, washed, and incubated with a metal-tagged anti-rabbit secondary antibody for 2 h at room temperature. After washing, sections were incubated with the metal-tagged panel overnight at 4 °C, stained with 0.5 μM Cell-ID Intercalator-Ir (Standard BioTools) for 7 min, washed three times in TBS for 10 min, rinsed in water, and air-dried.

#### Image acquisition

Images were acquired on a Hyperion XTI Imaging System (Standard BioTools). Each core was laser-ablated at a frequency of 800 Hz. Cores were selected for acquisition in random order, and each TMA was acquired in one continuous run.

#### Image processing, segmentation, and quality control

Raw data were processed with steinbock (v0.16.1) following the published IMC workflow (Windhager et al., 2023). MCD files were converted to OME-TIFF, and cells were segmented with the pre-trained Mesmer model (Greenwald et al., 2022) using nuclear (Ir191 and Ir193) and cytoplasmic (S100b, MelanA, SMA, CD31, CD45, and CD68) channels. Whole-cell masks containing zero or more than one nucleus were removed, and nuclei outside a whole-cell mask were expanded by 2 μm to generate cells. Mean marker intensities, morphological features (area and eccentricity), and neighbourhood information were extracted per cell, the last from a spatial cell graph constructed by pixel expansion in steinbock, as previously described (Meyer et al., 2026; Windhager et al., 2023). Channel spillover was compensated using a spillover slide (Chevrier et al., 2018). Single-cell data and images were imported into R with imcRtools and cytomapper (Eling et al., 2020) and stored as SingleCellExperiment objects (Amezquita et al., 2020). Mean intensities were arcsinh transformed with a cofactor of 1 (arc = asinh[raw]). Quality control was performed using pre-established criteria. At the cell level, objects with a segmented area of 5 μm^2^ or less were discarded as segmentation debris. At the image level, control cores containing normal tissue and cores with insufficient tissue or retraction artefacts were excluded following visual inspection. For panel I, 427 images from 52 patients (2,870,352 cells) were retained; for panel II, 412 images from the same 52 patients (2,513,554 cells). Marker performance was assessed per channel by fitting a two-component Gaussian mixture (mclust (Scrucca et al., 2016)) to the arcsinh values and taking the ratio of the mean raw intensities of the positive and negative components as the signal-to-noise ratio; a ratio of 2 was used as the reference line for flagging weak channels. Remaining ROIs and patients were then screened for outliers without further automatic exclusion: ROIs whose mean cell area or whose mean expression of any marker deviated from the cohort median by more than three median absolute deviations, and patients contributing fewer than two ROIs were flagged for manual review. Batch effects were examined by comparing marker distributions, ROI-level principal components, and silhouette widths across the three TMAs, and cell density and nearest-neighbour distance distributions were inspected per ROI.

#### Cell type classification

Cell type classification was performed with a customized version of an image-based neural network approach (CellSighter, **Fig. S3A**) (Amitay et al., 2023). A training set was created by labelling a subset of all detected cells by manual gating: cells that could be unambiguously assigned to one of eight broad types (tumour cells, vessels, stromal cells, CD4^+^ T cells, CD8^+^ T cells, myeloid cells, B cells, and plasma cells) on the basis of key lineage markers were added to the training set, and all other cells were labelled ‘undefined’ (**Fig. S3B**). A network was trained and tested on this dataset with an 80/20 split (**Fig. S3D**), and the trained network was then used to predict the type of every cell that had not been manually labelled. To confirm that the phenotype of network-labelled cells was consistent with that of manually labelled cells, the expression patterns of the two subsets were compared visually (**Fig. S3C, F**). The weighted F1 score of the network trained on manually labelled cells was 0.988 on the held-out test set. This evaluation systematically overestimates performance on the full dataset, because manually labelled cells are easier to classify by construction. To obtain a better estimate of performance on the full dataset, a second network was trained and tested using both manually labelled and network-labelled cells as ground truth; this quantifies the internal consistency of the classification used in all downstream analyses. As expected, the performance was lower, but the weighted F1 score remained above 0.9 (**Fig. S3G**). Classification was finally assessed by visual inspection of the classified images. The same approach was applied to panel II (**Fig. S3H–M**).

#### Immune cell state clustering/metaclustering

CD4^+^ T cells, CD8^+^ T cells, and myeloid cells were clustered separately on lineage-specific state markers: CD4^+^ T cells on TCF1/7, CTLA-4, LAG-3, PD-1, FOXP3, 4-1BB, Ki-67, and HLA-DR; CD8^+^ T cells on granzyme B, TCF1/7, CTLA-4, LAG-3, PD-1, 4-1BB, and Ki-67; and myeloid cells on CD15, IDO1, CD68, arginase-1, CD163, CD11c, PD-L1, and HLA-DR. Each lineage was clustered in two steps. In the first step, cells were clustered on their state markers with Rphenoannoy v0.1.0, a PhenoGraph implementation that builds a Jaccard-weighted nearest-neighbour graph and partitions it by Louvain community detection (Blondel et al., 2008; Levine et al., 2015). The number of nearest neighbours was swept over k = 50, 70, 90, 110, 130, and 160, and the resulting partitions were compared by cluster number, root-mean-square deviation of the cluster mean profiles, and silhouette width. The same value, k = 110, was used for all three lineages. This gave 23 clusters for CD4^+^ T cells, 18 for CD8^+^ T cells, and 19 for myeloid cells. In the second step, each first-step cluster was summarized by its mean arcsinh expression over the markers used for that lineage, and these cluster profiles were metaclustered by k-means. The number of metaclusters was chosen separately for each lineage over a range of 2 to 20 by internal cluster validation (clValid, Euclidean distance; connectivity, Dunn index, and silhouette width), with silhouette width as criterion. This gave 4 CD4^+^ T cell metaclusters, 5 CD8^+^ T cell metaclusters, and 5 myeloid metaclusters, which were then annotated by their marker expression and are the immune cell states used throughout. For the melanoma comparison, immune cells from both cohorts were clustered jointly by the same two-step procedure and annotated with the same scheme, giving 19 immune cell states.

#### Tumour cell clustering/metaclustering

Tumour cell phenotypes were defined in two steps, following the approach used previously (Meyer et al., 2026). First, tumour cells were clustered on tumour-compartment markers with the PhenoGraph algorithm (Jaccard index-based weights, Louvain algorithm for community detection) (Blondel et al., 2008; Levine et al., 2015), yielding the 34 tumour clusters. Clustering used the markers shown in Fig. 3A (CDK4, β-catenin, NGFR, HLA-DR, TYRP1, caveolin-1, HLA-ABC, mTOR, PD-L1, CA9, CDK6 and Ki-67), with k = 110 selected from a sweep of the neighbourhood size (**Fig. S8A**). Second, these fine clusters were grouped into tumour metaclusters (MCs) using CLARA clustering (Kaufman & Rousseeuw, 1990) with k = 5 on *z*-scored arcsinh expression of Ki-67, HLA-DR, NGFR, and TYRP1 resulting in 5 tumour metaclusters. In the validation cohort, tumour cells were clustered with the same procedure at k = 70, and the resulting clusters were assigned to MC1–MC5 by their defining markers.

#### ADC target scoring

Five criteria were computed per antigen on tumour cells: (1) median arcsinh expression; (2) percentage of positive tumour cells; (3) patient coverage, the percentage of patients with at least 25% positive tumour cells (sensitivity analyses at 10% and 50%); (4) homogeneity, the inverse coefficient of variation of raw tumour-cell intensity; and (5) selectivity, the median expression in tumour cells minus that in non-tumour cells. Positivity thresholds were set per marker where the weighted densities of a two-component Gaussian mixture (mclust (Scrucca et al., 2016)) crossed. Five markers (FOLR1, claudin-6, TROP2, NaPi2b, and EGFR) were scored 0 for positivity-based criteria. Two further antigens, cadherin-6 and tissue factor, had positive cells in fewer than 1% of tumour cells and no patient reaching the 25% coverage threshold, so seven of the fifteen antigens contribute essentially no positive signal. Each criterion was min-max scaled across all 15 antigens, pooled over both panels, and the composite score was their unweighted mean. Cross-panel comparisons used the cores present in both panels.

### BULK RNA SEQUENCING

Total RNA was extracted from frozen tissue with the Direct-zol RNA kit (Zymo Research), including on-column DNase digestion, and assessed by NanoDrop, Qubit (Thermo Fisher Scientific), and TapeStation (Agilent). Samples with an RNA integrity number above 6 and more than 250 ng RNA were used for library preparation with the KAPA Stranded mRNA-Seq kit (Roche). Libraries were sequenced on an Illumina HiSeq 2500 (paired-end 100 bp; about 60 million read pairs per library). Reads were aligned to GRCh37 with HISAT2 (Kim et al., 2019) and quantified with StringTie (Pertea et al., 2015). Immune cell fractions were obtained from the TIMER2.0 resource (Li et al., 2020), which applies several published deconvolution methods to a submitted expression matrix; the CIBERSORT (Chen et al., 2018; Newman et al., 2015) estimates of that output were used, for 23 primary tumour samples from 23 patients. Seven lineages (B cells, CD8^+^ T cells, CD4^+^ T cells, macrophages and dendritic cells, mast cells, eosinophils, and neutrophils) were formed by summing the corresponding signatures.

### DEEP VISUAL PROTEOMICS

DVP (Mund et al., 2022) was performed on tumour cores from 22 patients, selected by H&E-guided review of tumour regions and to represent short and long overall survival; the 21 analysed patients comprised 13 with short (2–36 months) and 8 with long (41–240 months) overall survival. A separate pilot acquisition (42 cores from four patients: 26 tumour, 16 stroma; **Fig. S10A–G**) is not part of the 164 cores below. Of the 164 cores acquired, 141 from 21 patients passed core quality control (**Sup. Table S6**); one patient (all nine of whose cores were excluded) therefore does not appear in any downstream analysis.

#### Laser microdissection

Sections (7 μm) of the FFPE TMAs (1-mm cores) were mounted on PPS frame slides (4-μm membrane, steel frame; Leica, 11600294), air-dried at room temperature, deparaffinized, rehydrated, stained with haematoxylin and eosin (H&E), and coverslipped with a xylene-based mounting medium. Reference marks were set on the membrane without perforating it (Leica LMD7 with HC PL Fluotar 10×/0.32 PH1 objective; laser power 15, aperture 1, speed 10, head current 100%, pulse frequency 528) to allow contours to be transferred between the digital annotation and the microdissection microscope. Slides were scanned on a Zeiss AxioScan at 20× magnification; the coverslips were then removed by gentle agitation in xylene (10 min, room temperature), residual mounting medium was washed off, and the slides were dipped in 100% ethanol, dried at 45 °C for 5 min, and stored at room temperature until microdissection. Tumour regions were annotated manually in QuPath (v0.5.1) by board-certified pathologists (N.D. and S.H.), covering a cumulative area of 20,000–25,000 μm² per tumour. Contours and reference marks were exported as GeoJSON and converted to the LMD7 XML format with a custom Python script. Contours were excised on a Leica LMD7 (LMD software v8.5.0.9136) with a 20× objective (HC PL FLUOTAR 20×/0.40 CORR) in brightfield mode (power 36–40, aperture 1, speed 11, middle pulse count 2, final pulse 4, head current 100%, pulse frequency 555, offset 119) into twin.tec 384-well LoBind PCR plates (Eppendorf, 0030129547) held at 32 °C. Contours were collected at the bottom of each well with 12 μL of 100% acetonitrile to rinse the well walls and vacuum-dried in a SpeedVac (60 °C, 20 min), and every well was inspected microscopically before sample preparation. The workflow followed, in principle, the published deep visual proteomics protocol (Mund et al., 2022; Nordmann et al., 2024). Sample preparation followed the ultra-low-input spatial tissue proteomics framework (Makhmut et al., 2023). For sample preparation, plates were sealed with adhesive PCR sealing foil (Thermo Fisher, AB-0626) and the tissue was incubated in 4 μL of 60 mM triethylammonium bicarbonate, 0.1% (w/v) n-dodecyl-β-D-maltoside, 5 mM TCEP, and 20 mM chloroacetamide at 95 °C for 60 min in a PCR cycler. After addition of 1 μL of 60% acetonitrile (final concentration 12%, v/v), samples were incubated at 75 °C for a further 60 min and digested with trypsin and LysC (4 ng/μL each) at 37 °C for 12 h. Peptides were vacuum-dried in a SpeedVac (60 °C, 20 min), resuspended in 20 μL of 2% acetonitrile and 0.1% trifluoroacetic acid containing custom iRT peptides (4 fmol each), and loaded onto C18 Evotips (Evotip Pure, Evosep, EV2011) according to the manufacturer’s instructions: tips were conditioned in solvent B (0.1% formic acid in acetonitrile), activated in isopropanol, equilibrated in solvent A (0.1% formic acid in water), loaded and washed once with solvent A, with every centrifugation at 800 g for 1 min; loaded tips were topped with solvent A and stored at 4 °C until analysis (Nordmann et al., 2024).

#### LC–MS/MS analysis

Peptides were separated on an Evosep One liquid chromatography system (Evosep Biosystems) with an Aurora Rapid 5 × 75 XT C18 column (IonOpticks), using the predefined 80 samples-per-day Whisper Zoom method (16.2-min gradient; solvent A, 0.1% formic acid in water; solvent B, 0.1% formic acid in acetonitrile), and analysed on an Orbitrap Astral mass spectrometer (Thermo Fisher Scientific) in data-independent acquisition (DIA) mode. MS1 spectra were acquired in the Orbitrap at a resolution of 240,000 (maximum injection time 5 ms; normalized AGC target 300%, corresponding to 3,000,000 charges). MS2 spectra were acquired in the Astral analyzer from 299 sequential, non-overlapping 2-m/z isolation windows over 380–980 m/z with optimized window placement, with higher-energy collisional dissociation at a normalized collision energy of 25%, a fragment range of 150–2,000 m/z, a scan time of 3 ms, a normalized AGC target of 300% (30,000 charges), and one microscan; multiplexed ion acquisition was disabled.

#### Raw data analysis with Spectronaut

Raw spectral data were analysed with Spectronaut (Biognosys, analysis version 20.5.260227) in library-free directDIA mode against the human Swiss-Prot FASTA database (release November 2021; 20,360 entries). Carbamidomethylation of cysteine was set as a fixed modification, and protein N-terminal acetylation and methionine oxidation as variable modifications; enzyme specificity was trypsin and LysC with up to two missed cleavages, and the minimum peptide length was seven amino acids. Proteins were inferred with ID Picker and quantified with the automated label-free quantification settings. The resulting library comprised 107,007 precursors, 95,773 peptides, and 7,365 protein groups (7,452 proteins). Across the 164 runs the mean number of missed cleavages was 0.29 (25.8% of peptides with at least one). The mean absolute mass deviations were 5.35 ppm at the MS1 level and 2.72 ppm at the MS2 level, and the median chromatographic peak width was 0.074 min. The median identification depth per core was 35,708 precursors, 33,326 peptides, and 5,388 protein groups, with the number of identified protein groups ranging from 2,066 to 6,172 per core before filtering.

#### Filtering approach per analysis

Data were processed in R 4.5.1, with no inter-sample normalization applied beyond the normalization performed during Spectronaut processing. A core was removed when its number of quantified protein groups fell more than 1.6 standard deviations below the median of its patient-by-region identifier, and nine cores, all from one patient, were removed on manual review. Within each identifier with at least two cores, a protein value was kept only when the protein was quantified in at least two-thirds of that identifier’s cores (the two-thirds rule), and identifier-level values were the median over the cores. After contaminant removal and core quality control, 7,276 protein groups in 141 cores from 21 patients entered the three functional analyses. Each analysis then applied its own filters, and the numbers of protein groups surviving each step are given below.

*Tumour centre versus invasive front* used 117 cores from 42 patient-by-region identifiers in 21 patients, with the two-thirds rule applied to identifiers contributing at least two cores. Starting from 7,276 protein groups, removing the matrisome, immune, epidermal, and adjacent-tissue compartments left 7,057 protein groups.

*Primary tumour versus lymph-node metastasis* used 47 cores from 18 identifiers in 9 patients, with the same at-least-two-core rule. Starting from 7,276 protein groups, removing the lymphoid and matrisome compartments left 7,181 protein groups.

*Short versus long survival* used 117 cores from 42 identifiers in 21 patients, with the two-thirds rule applied to groups of at least three cores. No compartment filter was applied to this analysis. Of the 7,276 protein groups, 6,917 met the estimability requirement of at least four observed values overall and at least two per survival group.

### H&E IMAGING AND DEEP LEARNING

FFPE sections (2 μm) consecutive to the IMC sections were deparaffinized and rehydrated as above, stained with haematoxylin (Dako, Agilent) for 3 min, blued in Scott’s tap water, counterstained with eosin Y (Dako) for 2 min, dehydrated, cleared in xylene, and mounted in Eukitt. Slides were scanned on an Aperio AT2 (Leica Biosystems) at 20× magnification (about 0.5 μm per pixel).

The analysis set comprised 194 tumour-centre core images from 43 patients (**Sup. Table S7**). The MC3 target was each patient’s MC3 fraction of tumour cells, dichotomized at the median of the 47-patient survival set, so that MC3-low and MC3-high denote the same patients as in Fig. 3H; the Ki-67 target was the per-patient mean tumour-cell Ki-67, dichotomized at its median in the same set. The two labels agree for 41 of the 43 patients. Patient labels were assigned to all cores of that patient. Following the TRIDENT workflow (Zhang et al., 2025), cores were tiled into non-overlapping 224 × 224-pixel patches; a patch was retained if at least 50% of its pixels had a greyscale intensity below 220, and at most 200 patches were sampled per core. Patches were embedded with Phikon-v2 (Filiot et al., 2024) (1,024 dimensions). For morphological curation, embeddings were clustered by k-means (k = 10; 10 initializations; maximum 300 iterations); one cluster of predominantly non-tumour morphology was excluded after pathologist review, leaving 34,558 of 38,557 patches.

For the SVM (Cortes & Vapnik, 1995) (radial basis function kernel) and random forest (Breiman, 2001) (200 trees), patch embeddings of each core were summarized by their mean and maximum, concatenated (2,048 dimensions), and standardized; both used balanced class weights. The Attn-MIL model (Ilse et al., 2018) mapped each patch embedding through a 128-dimensional attention branch (tanh, dropout 0.3) to a normalized attention weight, and the attention-weighted core representation was passed to a classifier with one 128-dimensional hidden layer (ReLU, dropout 0.3). It was trained for 30 epochs with Adam (Kingma & Ba, 2014) (learning rate 10^−3^, weight decay 10^−4^), class-weighted binary cross-entropy, and gradient clipping at norm 1.0. Models were evaluated by stratified five-fold cross-validation grouped by patient (StratifiedGroupKFold). UMAP (McInnes et al., 2018) of up to 8,000 sampled patch embeddings (15 neighbours, minimum distance 0.3) was used for visualization. Models were implemented with scikit-learn (Pedregosa et al., 2012) and PyTorch (Paszke et al., 2019).

### IMMUNOHISTOCHEMISTRY

FFPE sections were cut onto adhesive slides, dried, deparaffinized in xylene and rehydrated through a graded ethanol series. Heat-induced epitope retrieval was performed, and endogenous peroxidase activity was blocked with hydrogen peroxide. Sections were incubated with primary antibodies against c-MET (Abcam, ab227637) and B7-H3 (Abcam, ab105922), followed by a horseradish-peroxidase-conjugated polymer detection system. The signal was developed with 3,3’-diaminobenzidine, and sections were counterstained with haematoxylin (Dako, Agilent), dehydrated, cleared in xylene, and mounted. Stained slides were scanned and evaluated by pathologists (S.H. and C.P.).

### STATISTICAL ANALYSIS

#### General principles

The unit of analysis for all between-region, between-cohort, and survival comparisons was the patient: cell- and ROI-level values were aggregated per patient (and per region where applicable) before testing. The numbers of cells, ROIs, and patients underlying each panel, the statistical test, the centre and dispersion shown, and the multiple-testing family are given in the figure legends. Tests were two-sided. Multiple testing was corrected with the Benjamini-Hochberg (BH) method (Benjamini & Hochberg, 1995) within the families named in each legend; edgeR analyses were called at a false discovery rate (FDR) below 0.1 and all other analyses at 0.05. Given the cohort size, all analyses are exploratory, and P values are interpreted as evidence of association rather than confirmation.

#### Composition, density, and diversity

Cell-type fractions were computed per ROI among all cells or among immune/tumour cells. Densities (cells/mm^2^) used an assumed ROI area of 0.785 mm^2^ (the area of a 1-mm core) for CCS and 1.0 mm^2^ for melanoma. Diversity was quantified per ROI by Shannon entropy (Shannon, 1948), *H* = −Σ *p*_i_ ln *p*_i_, over cell-type proportions and averaged per patient; Gini-Simpson index and Pielou’s evenness (*H*/ln *S*) were also computed. Paired region comparisons used the Wilcoxon signed-rank test on patient-level means in patients with both regions (40 patients); cohort comparisons used the Wilcoxon rank-sum test on patient-level values. Tumour MC co-occurrence (**Fig. S8F–G**) was assessed using MC fractions at the ROI and patient levels. Zero fractions were replaced with half the smallest positive fraction in the matrix before centred log-ratio transformation. Spearman correlations were then calculated for all ten MC pairs. To account for the dependence introduced by fractions summing to one, correlations were compared with a permutation null model: each MC’s fractions were independently shuffled across ROIs or patients, rows were rescaled to sum to one, and the transformation and correlations were recalculated. Z-scores were then calculated to show how far each observed correlation was above or below the mean correlation expected under the null model.

#### Differential abundance

Cell counts were summed per patient and region, normalized by TMM (Robinson & Oshlack, 2010), and tested with edgeR (Chen et al., 2025; Robinson et al., 2010) quasi-likelihood F-tests (design ∼patient + region; dispersion estimated without trend; robust quasi-likelihood fit). The same approach was used for primary tumour versus metastasis (∼patient + site) and for CCS versus melanoma (∼cohort, on patient-summed counts).

#### Spatial interaction analysis

Tumour-partner interaction counts were computed with imcRtools (countInteractions, method “classic”) on the precomputed neighbourhood graph (Meyer et al., 2026; Windhager et al., 2023). For each ROI and each ordered pair (tumour → target) this returns the contact rate ct, the number of observed directed tumour → target edges divided by the number of tumour cells in that ROI (**Fig. S5A**). Tumour metaclusters were collapsed to a single tumour label. To compare tumour–macrophage engagement between regions independently of abundance, the contact rate was modelled across ROIs by ordinary least squares as ct ∼ target-cell frequency + tumour-cell frequency + cell density, where cell density is the mean number of spatial neighbours per cell in the ROI (mean undirected degree). The covariates are shown in **Fig. S5B** and **Fig. S5C**. One model was fitted per target population, within the analysis set (**Fig. S5D**; adjusted R² = 0.712, 0.881 and 0.692), and the infiltration score was defined as log2(ct / predicted ct) (**Fig. S5E**). Regions were compared with paired Wilcoxon signed-rank tests on the per-patient scores, the patient being the unit of analysis, with Benjamini-Hochberg correction across the three macrophage populations.

#### Survival analysis

##### Outcome definition

Overall survival (OS) was defined with death from disease as the event; patients recorded as alive with or without disease were censored. The event indicator was constructed from the status (death from disease = 1; alive with no evidence of disease or alive with disease = 0). Survival time in days and status were extracted from the clinical data. Median follow-up, estimated by the reverse Kaplan-Meier method, was 81 months (2,465 days; 47 patients). Hazard ratios were estimated with Cox proportional-hazards models (Cox, 1972) using the survival package, and Kaplan-Meier curves were compared by log-rank tests (survminer). The proportional-hazards assumption of the reported models was assessed with scaled Schoenfeld residuals (Grambsch-Therneau test) (Grambsch & Therneau, 1994).

##### Tumour metacluster analysis

The analysis set comprised patients with at least 50 tumour cells and usable follow-up. Of the 52 patients analysed by IMC, three contributed only lymph-node metastasis ROIs and therefore have no primary-tumour metacluster composition; of the remaining 49, two were excluded for missing or non-positive survival time and none by the cell-count floor, leaving 47 patients with 35 events. Each MC fraction was dichotomized at the cohort median and, separately, at the cohort mean; both cut-points were specified in advance and both are reported. Five separate models were fitted per cut-point, one per MC, each adjusted for age (per 10 years), metastatic status at diagnosis, and sex. BH correction was applied over the five MC terms within each cut-point. As a sensitivity analysis without a cut-point, the MC3 fraction was also entered as a continuous variable (hazard ratio per 10 percentage points) in a Cox model with the same covariates, in the same 47 patients with 35 events. Per-patient Ki-67 was the mean of min-max-scaled arcsinh Ki-67 over tumour cells, dichotomized at the median.

##### ADC target expression

c-MET and B7-H3 were summarized per patient as the mean arcsinh expression over tumour cells in primary-site cores, with lymph-node metastases excluded, and dichotomized at the cohort median. The cut-point is the median and was prespecified. The reported P value is the log-rank P, BH-corrected.

##### Proteomic module scores

Patients were first assigned to a short-survival or a long-survival group. A module score was then computed for each protein module and patient. Each protein was standardized across patients, and the score is the mean of those standardized values over the proteins of the module. Only modules with at least 10 proteins quantified in at least 70% of patients were scored, which left 47 modules. Each module score was tested against overall survival as a continuous variable (per standard deviation) in a separate Cox model. Because the modules shown were selected on the same patients, family-wise error was assessed by permutation: the survival outcome was permuted across patients 2,000 times, the maximum absolute Cox z statistic over the 47 modules was recorded for each permutation, and p_FWER is the fraction of permutations in which that maximum reached the observed |z|.

##### External validation cohort

This cohort contributed 13 patients with usable follow-up, among whom 10 died. With 10 events no covariate can be estimated alongside the variable of interest, so all models in this cohort are unadjusted and the cut-point is the cohort median, as in the discovery cohort.

##### Interpretation

The discovery cohort comprises 52 patients, of whom 47 enter the survival analysis with 35 events, and the validation cohort 13 patients with 10 events. With cohorts of this size the survival results are therefore exploratory and hypothesis-generating.

#### Mass spectrometry proteomics statistics

Differential protein abundance was tested with limma (Ritchie et al., 2015) at core level with patient as a blocking factor via duplicateCorrelation (Smyth et al., 2005), requiring at least four observed values overall and two per group; BH correction was applied over all tested proteins with thresholds of FDR < 0.05 and |log_2_ fold change| ≥ 1. Gene-set enrichment used fgsea (Korotkevich et al., 2016) on the moderated t statistic (the most extreme value per gene symbol) with the MSigDB Hallmark collection (Liberzon et al., 2015) (minimum size 10, maximum size 500). For tumour centre versus invasive front (**Fig. 4C**), proteins were ranked by a paired moderated t statistic (limma, ∼patient + region, on patient-by-region median profiles) and tested against 28 curated sets. For primary tumour versus metastasis (**Fig. 6**), the core-level test was complemented by a patient-level test: the identifier-level values were differenced within each patient (metastasis minus primary), and a moderated one-sample t test (limma) was applied to proteins with a difference in at least three patients, with BH correction over all proteins tested. Gene-set enrichment for this comparison used 21 gene-sets (tumour-intrinsic programmes, therapeutically targetable axes, and neural crest development), ranked on the core-level moderated t statistic.

#### Primary tumour versus metastasis

MC counts were converted to centred log-ratios (log2[(count + 1)/total], centred per sample) (Aitchison, 1982). Composition was compared by PERMANOVA (Anderson, 2001) on Euclidean distances of centred log-ratio values (Aitchison distance) with the adonis2 function of the vegan R package (Oksanen et al., 2026). Individual MCs were compared by paired Wilcoxon signed-rank tests with BH over five MCs and by edgeR as above.

#### Classifier evaluation

Performance was summarized from out-of-fold predictions by AUC-ROC, average precision, weighted F1 score, Matthews correlation coefficient, and confusion matrices at a probability threshold of 0.5. Out-of-fold core probabilities were averaged per patient to give the patient-level scores reported in the Results, since cores from one patient are not independent. Confidence intervals on the patient-level AUC-ROC are percentile bootstrap intervals over 2,000 resamples of patients, and significance was assessed against a null obtained by permuting the patient labels 2,000 times. No external validation cohort was available for the H&E models.

#### Software and package versions

For IMC, raw data were processed with steinbock v0.16.1 (Windhager et al., 2023). For deep visual proteomics, tumour regions were annotated in QuPath v0.5.1 and microdissected with the Leica LMD software v8.5.0.9136, and the raw data were analysed with Spectronaut v20.5.260227 (Biognosys). The H&E deep-learning analysis was run in Python v3.11.5 with TRIDENT (Zhang et al., 2025), the Phikon-v2 foundation model (Filiot et al., 2024), scikit-learn v1.8.0 (Pedregosa et al., 2012), PyTorch v2.10.0 (Paszke et al., 2019), umap-learn v0.5.11 (McInnes et al., 2018), NumPy v1.24.3, pandas v2.3.3, SciPy v1.11.1, and matplotlib v3.7.2.

Statistical analyses and figures were produced in R (R Core Team, 2026). Data preprocessing was performed in R v4.5.1, and downstream analyses and figure generation in R v4.6.2; the complete session information for both environments is provided in the GitHub repository (see Code availability). Analyses and visualizations were performed using the following R packages: SingleCellExperiment v1.34.0, imcRtools v1.18.1, cytomapper v1.24.0, scater v1.40.2, uwot v0.2.5, Rphenoannoy v0.1.0, cluster v2.1.8.3, clValid v0.7, mclust v6.1.3, edgeR v4.10.5, limma v3.68.5, fgsea v1.38.0, msigdbr v26.1.1, survival v3.8-12, survminer v0.5.2, vegan v2.7-6, ComplexHeatmap v2.28.0, circlize v0.4.18, dittoSeq v1.24.0, ggplot2 v4.0.3, and patchwork v1.3.2.

#### Code availability

All original code used for the analyses in this study has been deposited on GitHub and is publicly available as of the date of publication (https://github.com/BodenmillerGroup/CCS_project).

