## Supplementary Figures for "Spatial proteomics identifies a myeloid-dominated, compartmentalized microenvironment and a proliferation-associated prognostic signature in clear cell sarcoma"

**SUPPLEMENTAL INFORMATION**

Table of contents

1. FIGURES S1–S11

#### 1. FIGURES S1–S11

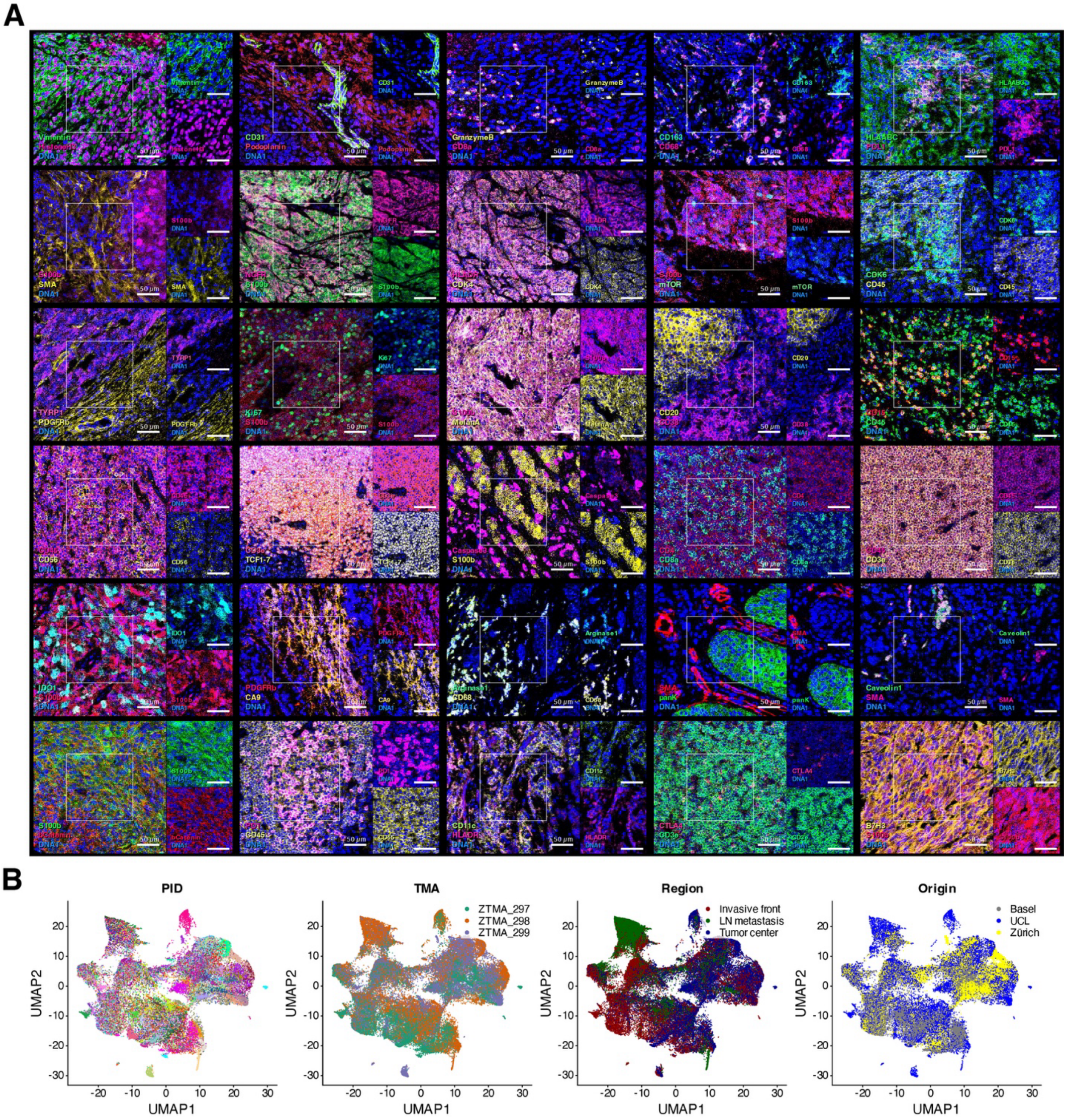


**Figure S1: Quality control of IMC antibody panel I.** *Related to **Figure 1**.*
**(A)** Representative multichannel IMC images showing staining with panel I antibodies.
**(B)** UMAP embedding computed on the broad phenotyping markers and shown four times, coloured by patient (PID), TMA of acquisition, ROI category and centre of origin.

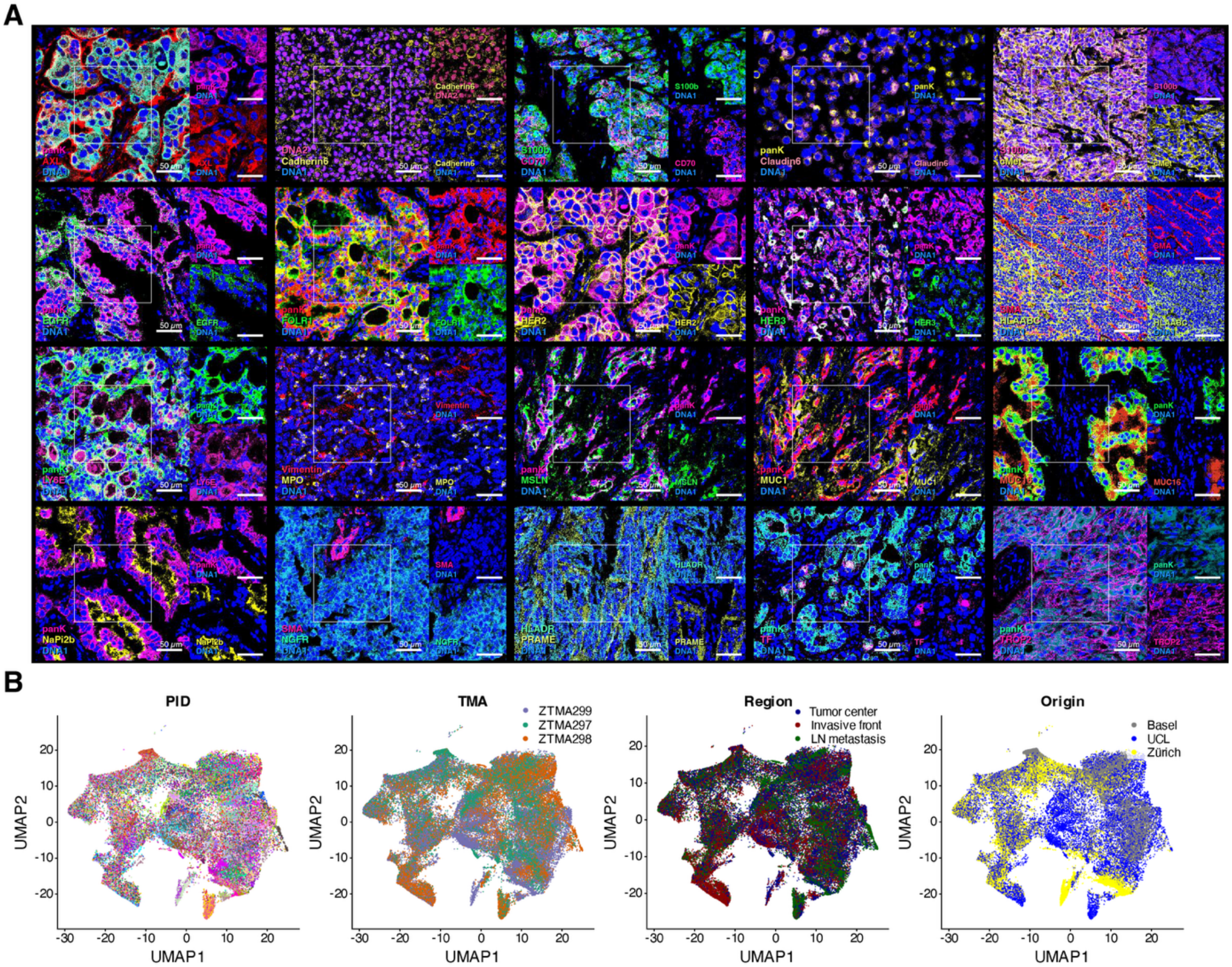


**Figure S2: Quality control of IMC antibody panel II (ADC targets).** *Related to **Figure 1**.*

**(A)** Representative multichannel IMC images showing staining with panel II antibodies in cohort samples and positive control tissues.

**(B)** UMAP embedding computed on the broad phenotyping markers and shown four times, coloured by patient (PID), TMA of acquisition, ROI category and centre of origin.

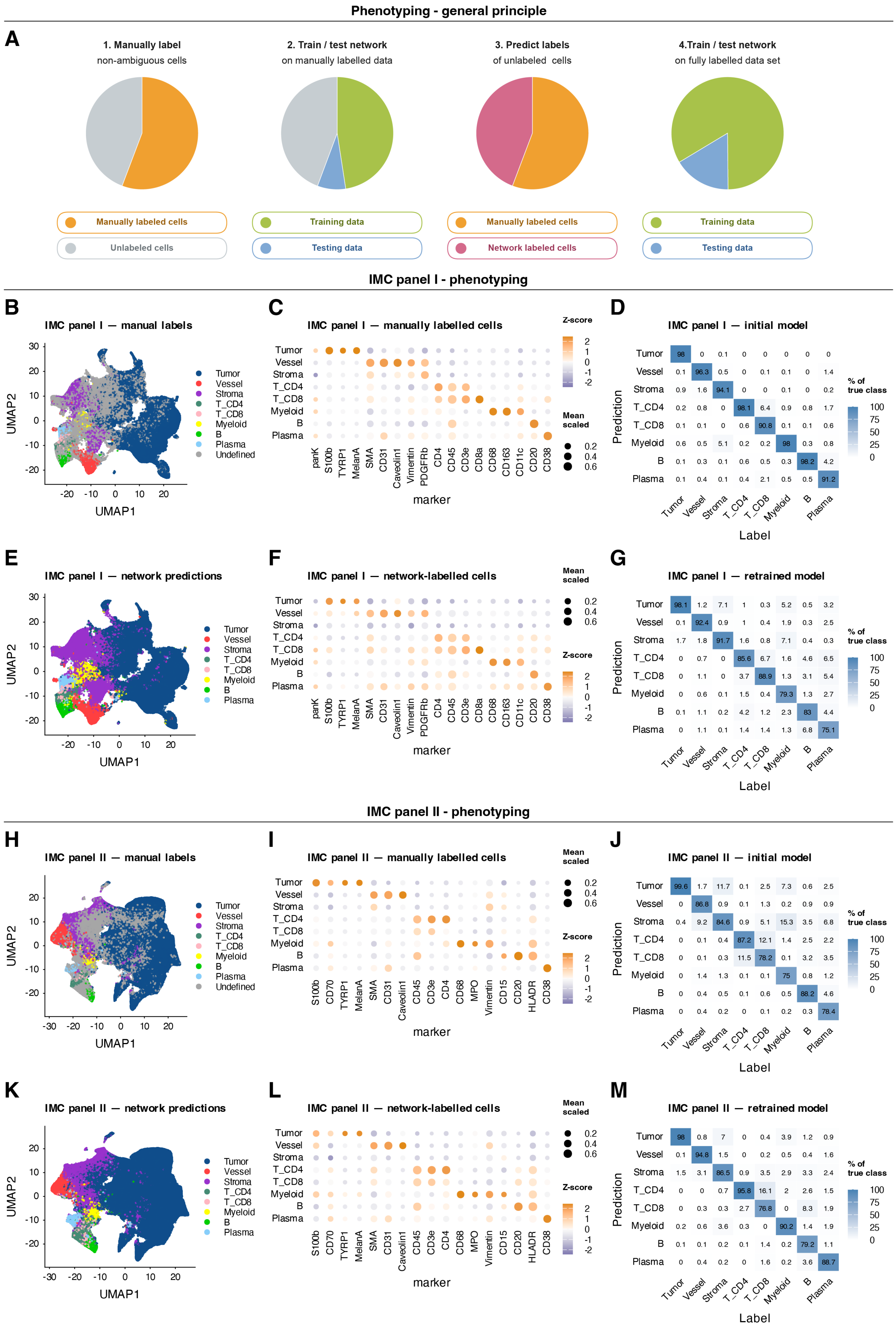
Phenotyping - general principle
A
1. Manually label
non-ambiguous cells
2. Train / test network
on manually labelled data
3. Predict labels
of unlabeled cells
4.Train / test network
on fully labelled data set
Manually labeled cells
Unlabeled cells
Training data
Testing data
Manually labeled cells
Network labeled cells
Training data
Testing data
IMC panel I - phenotyping
B
IMC panel I — manual labels
C
IMC panel I — manually labelled cells
D
IMC panel I — initial model
Tumor 98 0 0.1 0 0 0 0 0
Vessel 0.1 96.3 0.5 0 0.1 0.1 0 1.4
Stroma 0.9 1.6 94.1 0 0 0.1 0 0.2
T_CD4 0.2 0.8 0 98.1 6.4 0.9 0.8 1.7
T_CD8 0.1 0.1 0 0.6 90.8 0.1 0.1 0.6
Myeloid 0.6 0.5 5.1 0.2 0.2 98 0.3 0.8
B 0.1 0.3 0.1 0.6 0.5 0.3 98.2 4.2
Plasma 0.1 0.4 0.1 0.4 2.1 0.5 0.5 91.2
E
IMC panel I — network predictions
F
IMC panel I — network-labelled cells
G
IMC panel I — retrained model
Tumor 98.1 1.2 7.1 1 0.3 5.2 0.5 3.2
Vessel 0.1 92.4 0.9 1 0.4 1.9 0.3 2.5
Stroma 1.7 1.8 91.7 1.6 0.8 7.1 0.4 0.3
T_CD4 0 0.7 0 85.6 6.7 1.6 4.6 6.5
T_CD8 0 1.1 0 3.7 88.9 1.3 3.1 5.4
Myeloid 0 0.6 0.1 1.5 0.4 79.3 1.3 2.7
B 0.1 1.1 0.2 4.2 1.2 2.3 83 4.4
Plasma 0 1.1 0.1 1.4 1.4 1.3 6.8 75.1
IMC panel II - phenotyping
H
IMC panel II — manual labels
I
IMC panel II — manually labelled cells
J
IMC panel II — initial model
Tumor 99.6 1.7 11.7 0.1 2.5 7.3 0.6 2.5
Vessel 0 86.8 0.9 0.1 1.3 0.2 0.9 0.9
Stroma 0.4 9.2 84.6 0.9 5.1 15.3 3.5 6.8
T_CD4 0 0.1 0.4 87.2 12.1 1.4 2.5 2.2
T_CD8 0 0.1 0.3 11.5 78.2 0.1 3.2 3.5
Myeloid 0 1.4 1.3 0.1 0.1 75 0.8 1.2
B 0 0.4 0.5 0.1 0.6 0.5 88.2 4.6
Plasma 0 0.4 0.2 0 0.1 0.2 0.3 78.4
K
IMC panel II — network predictions
L
IMC panel II — network-labelled cells
M
IMC panel II — retrained model
Tumor 98 0.8 7 0 0.4 3.9 1.2 0.9
Vessel 0.1 94.8 1.5 0 0.2 0.5 0.4 1.6
Stroma 1.5 3.1 86.5 0.9 3.5 2.9 3.3 2.4
T_CD4 0 0 0.7 95.8 16.1 2 2.6 1.5
T_CD8 0 0.3 0.3 2.7 76.8 0 8.3 1.9
Myeloid 0.2 0.6 3.6 0.3 0 90.2 1.4 1.9
B 0.1 0.1 0.2 0.1 1.4 0.2 79.2 1.1
Plasma 0 0.4 0.2 0 1.6 0.2 3.6 88.7
UMAP1
UMAP2
Tumor
Vessel
Stroma
T_CD4
T_CD8
Myeloid
B
Plasma
Undefined
marker
Z-score
Mean scaled
Prediction
Label
% of true class

**Figure S3: CellSighter-based cell phenotyping for both antibody panels.** *Related to **Figures 1 and 7**.*
**(A)** Schematic of the four-step phenotyping approach: manual annotation of non-ambiguous cells, training and testing of the network on the manually labelled data, prediction of the labels of the unlabelled cells, and retraining of the network on the fully labelled data set. Pie chart areas are schematic and not to scale; they do not represent the relative proportions of cells or labels.
**(B–G)** Phenotyping of the data from IMC antibody panel I.
**(B)** UMAP of the manually labelled cells, coloured by cell type; the cells left Undefined by manual labelling are shown in grey. **(C)** Bubble plot of the mean scaled expression of the 18 markers per cell type among the manually labelled cells; dot size indicates the mean scaled expression and colour denotes the z-scored expression. **(D)** Confusion matrix of the initial model on held-out test cells. **(E)** The same UMAP as in B, coloured by network prediction. **(F)** Bubble plot as in C, for the cells left Undefined by manual labelling and assigned by the network. **(G)** Confusion matrix of the retrained model. Confusion matrices are normalized within each true class (columns) and are computed on held-out test cells; the weighted F1 score was 0.988 for the model trained on manually labelled cells and above 0.9 for the retrained model.
**(H–M)** The same six panels for IMC antibody panel II.

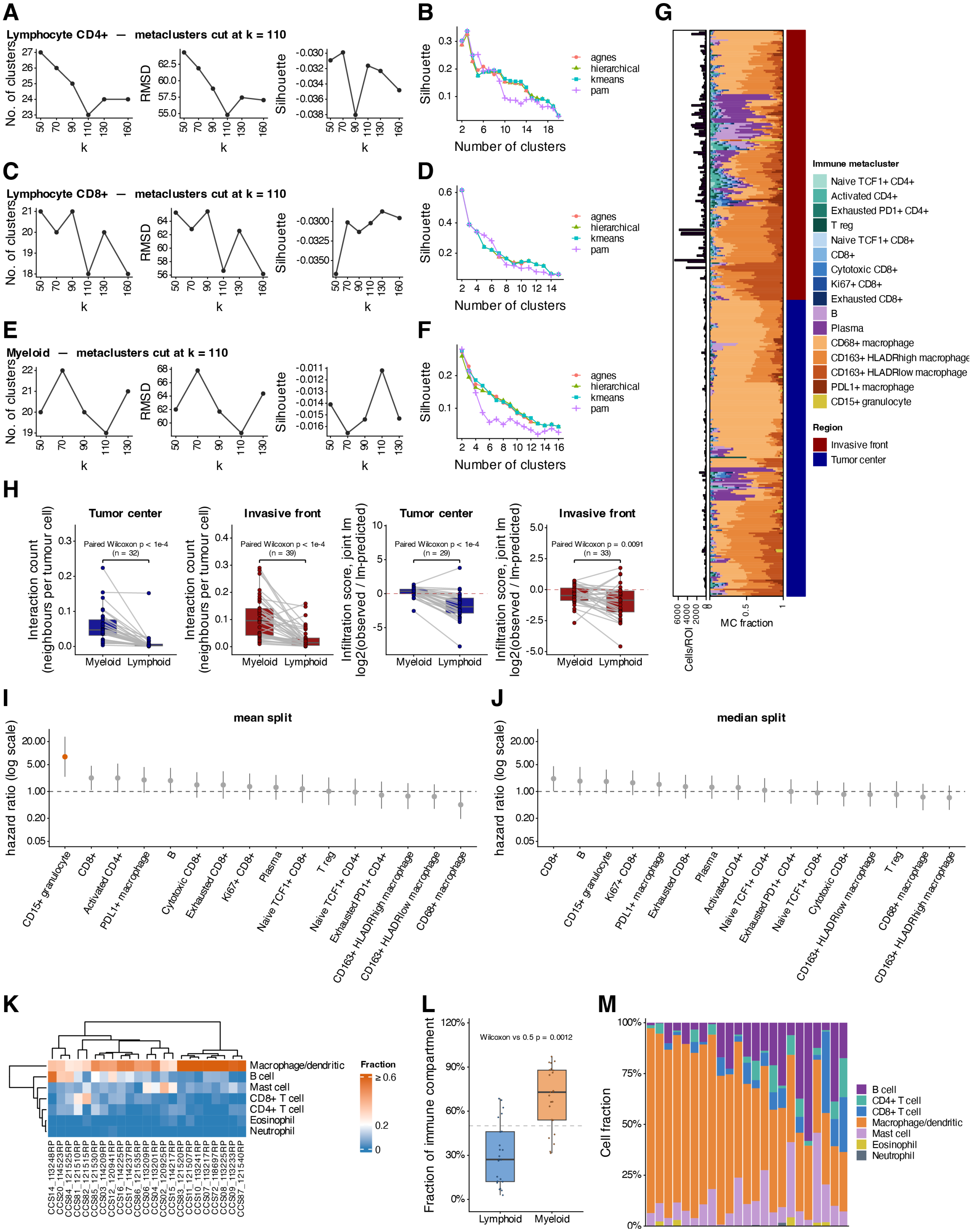


**Figure S4: Immune clustering, spatial immune analysis and association with survival.** *Related to* ***Figure 2.***

**(A, C, E)** Parameter selection for graph-based clustering of the CD4$^{+}$ T-cell (A), CD8$^{+}$ T-cell (C) and myeloid (E) compartments. The number of clusters (left), the root-mean-square deviation (RMSD; centre) and the silhouette width (right) are plotted against the PhenoGraph neighbourhood size k, the number of nearest neighbours used to build the cell graph. A k of 110 was selected for downstream analysis.

**(B, D, F)** Silhouette width for different numbers of clusters and four algorithms (hierarchical, agnes, k-means and PAM) for the $CD4^+$ T cells (B), the $CD8^+$ T cells (D), and the myeloid cells (F).

**(G)** Stacked bar plots of the per-ROI composition of the immune subtypes, ordered by the proportion of subtypes; the bar plot at left shows the number of cells per ROI and the ROI type is indicated as an annotation at right.

**(H)** Box plots of tumour-to-myeloid and tumour-to-lymphoid contacts in the tumour centre and the invasive front, shown both as the raw contact rate (neighbours per tumour cell) on the left and as the abundance-corrected infiltration score (log2 observed/lm-predicted contact rate; see Methods) on the right. Box plots show the median, interquartile range (IQR), and whiskers extend to the most extreme values within 1.5×IQR from the quartiles; each point is one patient and lines connect the two values of a patient. P values are indicated (paired Wilcoxon signed-rank test).

**(I, J)** Hazard ratios of overall survival estimated using Cox proportional-hazards models of survival adjusted for age, sex and metastatic status at diagnosis with abundance of immune population dichotomized at the mean (I) or at the median (J). Points, hazard ratio; bars, 95% confidence interval; BH correction. 47 patients with 35 events.

**(K)** Heatmap of the fractions of the indicated immune-lineage cells deconvolved (CIBERSORT) from bulk RNA-seq data (n = 23 samples). Rows and columns are hierarchically clustered.

**(L)** Box plots of the lymphoid and the myeloid fraction of the deconvolved immune signal from the data in K. Box plots show the median, interquartile range (IQR), and whiskers extend to the most extreme values within 1.5×IQR from the quartiles. P values were determined using a one-sample Wilcoxon signed-rank test against 0.5.

**(M)** Stacked bar plots of the per-sample lineage fractions from the bulk RNA-seq deconvolution in K.

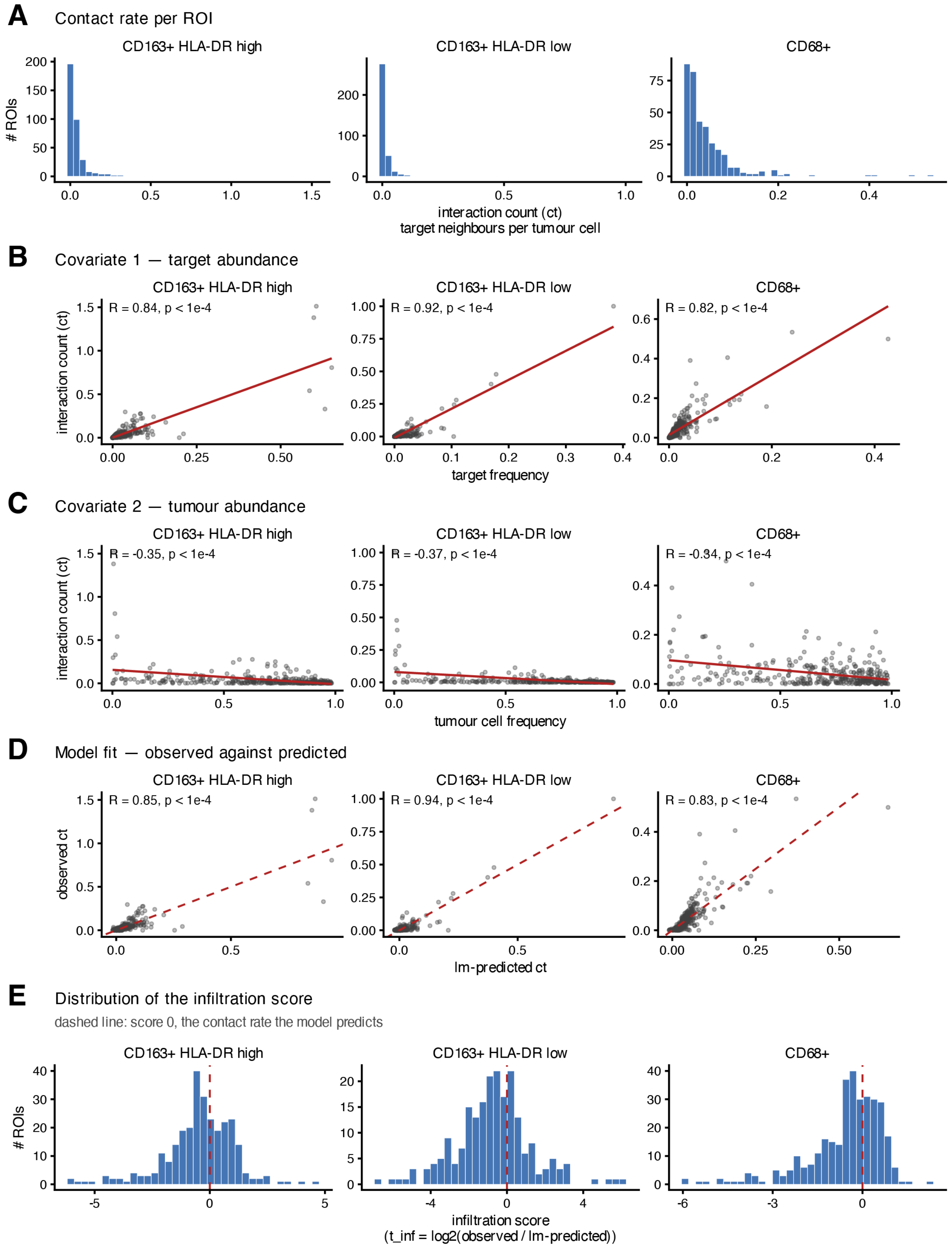
A
Contact rate per ROI
CD163+ HLA-DR high
CD163+ HLA-DR low
CD68+
### ROIs
interaction count (ct)
target neighbours per tumour cell
B
Covariate 1 — target abundance
R = 0.84, p < 1e-4
R = 0.92, p < 1e-4
R = 0.82, p < 1e-4
target frequency
C
Covariate 2 — tumour abundance
R = -0.35, p < 1e-4
R = -0.37, p < 1e-4
R = -0.34, p < 1e-4
tumour cell frequency
D
Model fit — observed against predicted
observed ct
R = 0.85, p < 1e-4
R = 0.94, p < 1e-4
R = 0.83, p < 1e-4
lm-predicted ct
E
Distribution of the infiltration score
dashed line: score 0, the contact rate the model predicts
infiltration score
(t_inf = log2(observed / lm-predicted))

**Figure S5: Components of the infiltration score used in the linear model.** *Related to* ***Figure 2****.*
Each panel is faceted by the three macrophage populations shown in Figure 2K ($CD163^+$ HLA-DRhigh, $CD163^+$ HLA-DRlow, $CD68^+$).
**(A)** Distribution of the contact rate *ct* per ROI, the number of observed directed tumour → target edges divided by the number of tumour cells in that ROI.
**(B, C)** *ct* against two covariates of the model, target-cell frequency and tumour-cell frequency. Red line, ordinary least-squares fit; *R*, Pearson correlation with corresponding P value.
**(D)** Observed *ct* against the contact rate predicted by the model, fitted separately for each target population within the analysis set (adjusted $R^2$ indicated); dashed line, identity.
**(E)** Distribution of the infiltration score, log2(*ct* / predicted *ct*), on the ROIs where it is defined. Dashed line, score 0, that is, the contact rate the model predicts.

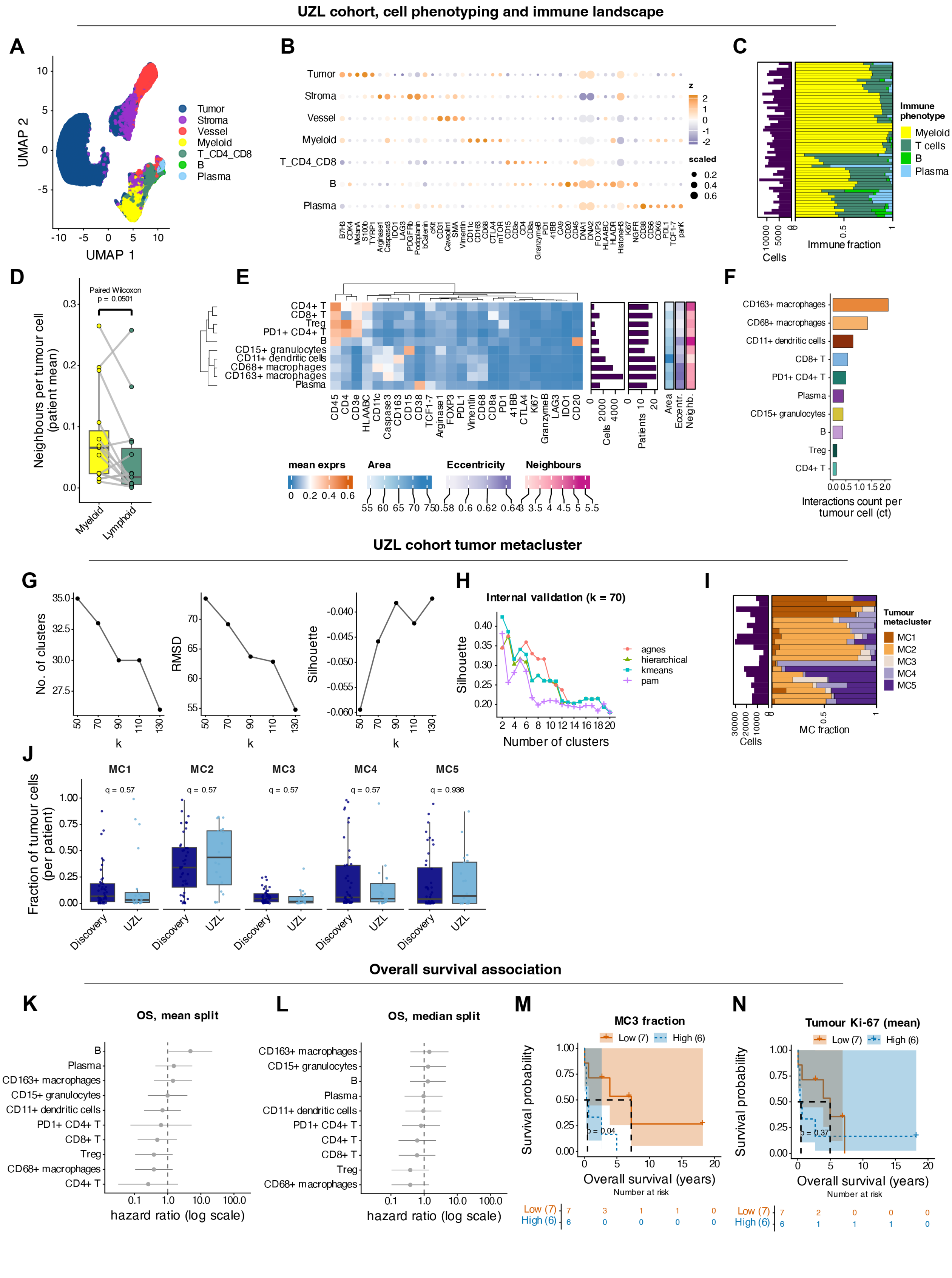
UZL cohort, cell phenotyping and immune landscape
A
UMAP 1
UMAP 2
Tumor
Stroma
Vessel
Myeloid
T_CD4_CD8
B
Plasma
B
Tumor
Stroma
Vessel
Myeloid
T_CD4_CD8
B
Plasma
z
scaled
C
Cells
Immune fraction
Immune phenotype
Myeloid
T cells
B
Plasma
D
Paired Wilcoxon
p = 0.0501
Neighbours per tumour cell (patient mean)
Myeloid
Lymphoid
E
CD4+ T
CD8+ T
Treg
PD1+ CD4+ T
B
CD15+ granulocytes
CD11+ dendritic cells
CD68+ macrophages
CD163+ macrophages
Plasma
Cells
Patients
Area
Eccentr.
Neighb.
mean exprs
Area
Eccentricity
Neighbours
F
CD163+ macrophages
CD68+ macrophages
CD11+ dendritic cells
CD8+ T
PD1+ CD4+ T
Plasma
CD15+ granulocytes
B
Treg
CD4+ T
Interactions count per tumour cell (ct)
UZL cohort tumor metacluster
G
No. of clusters
RMSD
Silhouette
k
H
Internal validation (k = 70)
Silhouette
agnes
hierarchical
kmeans
pam
Number of clusters
I
Cells
MC fraction
Tumour metacluster
MC1
MC2
MC3
MC4
MC5
J
MC1
MC2
MC3
MC4
MC5
q = 0.57
q = 0.57
q = 0.57
q = 0.57
q = 0.936
Fraction of tumour cells (per patient)
Discovery
UZL
Overall survival association
K
OS, mean split
B
Plasma
CD163+ macrophages
CD15+ granulocytes
CD11+ dendritic cells
PD1+ CD4+ T
CD8+ T
Treg
CD68+ macrophages
CD4+ T
hazard ratio (log scale)
L
OS, median split
CD163+ macrophages
CD15+ granulocytes
B
Plasma
CD11+ dendritic cells
PD1+ CD4+ T
CD4+ T
CD8+ T
Treg
CD68+ macrophages
hazard ratio (log scale)
M
MC3 fraction
Low (7)
High (6)
p = 0.04
Survival probability
Overall survival (years)
Number at risk
Low (7) 7 3 1 1 0
High (6) 6 0 0 0 0
N
Tumour Ki-67 (mean)
Low (7)
High (6)
p = 0.37
Survival probability
Overall survival (years)
Number at risk
Low (7) 7 2 0 0 0
High (6) 6 1 1 1 0

**Figure S6: Validation CCS cohort: immune landscape, tumour metaclusters and survival.** *Related to* ***Figures 2 and 3****.*

**(A)** UMAP of the single cells of the validation cohort (n = 20 patients; 10% downsampled for visualization), coloured by broad cell phenotype.

**(B)** Bubble plot of marker expression in the indicated cell types; dot size indicates the mean scaled expression of each marker and colour denotes the z-scored expression.

**(C)** Stacked bar plots of the per-ROI composition of the broad immune cell types in the validation cohort; the bar plot at left shows the number of cells per core.

**(D)** Box plot of tumour-to-myeloid and tumour-to-lymphoid contacts, shown as the raw contact rate (neighbours per tumour cell, patient mean). Box plots show the median, interquartile range (IQR), and whiskers extend to the most extreme values within 1.5×IQR from the quartiles; each point is one patient and lines connect the two values of a patient. P values are indicated (paired Wilcoxon signed-rank test).

**(E)** Heatmap of marker expression (z-score) per immune subtype, with the total number of cells and patients per subtype and the average spatial single-cell information (area, eccentricity and number of neighbours) shown alongside. Rows and columns are hierarchically clustered using Ward's method (ward.D2).

**(F)** Bar plot of the interaction count per tumour cell, from tumour cells to the indicated immune cell partners.

**(G)** Parameter selection for graph-based clustering of tumour cells. The number of clusters (left), the root-mean-square deviation (RMSD; centre) and the silhouette width (right) are plotted against the PhenoGraph neighbourhood size k, the number of nearest neighbours used to build the cell graph. A k of 70 was selected for downstream analysis.

**(H)** Silhouette width for different numbers of tumour-cell clusters and four algorithms (hierarchical, agnes, k-means and PAM). Each grouping is scored by its silhouette width.

**(I)** Stacked bar plots of the per-core composition of the tumour metaclusters (MCs), ordered by the proportion of MCs; the bar plot at left shows the number of tumour cells per core.

**(J)** Box plots of the fraction of each patient's tumour cells in each MC, in the discovery cohort and in the validation cohort. Box plots show the median, interquartile range (IQR), and whiskers extend to the most extreme values within 1.5×IQR from the quartiles; each point is one patient. P values are indicated (Wilcoxon rank-sum tests).

**(K, L)** Hazard ratios of overall survival estimated using univariable Cox proportional hazards models for each of the 10 immune subtypes, with abundance dichotomized at the mean (K) or at the median (L). Points show the hazard ratio and bars the 95% confidence interval, on a log scale. Data are from 13 patients with usable follow-up and 10 events; no covariate could be added to the models.

**(M, N)** Kaplan-Meier curves of overall survival for patients split at the median of the MC3 fraction (M) and of the mean tumour-cell Ki-67 expression (N), with number-at-risk tables; 13 patients (7 low, 6 high) and 10 events. The indicated P values were calculated using the log-rank test.

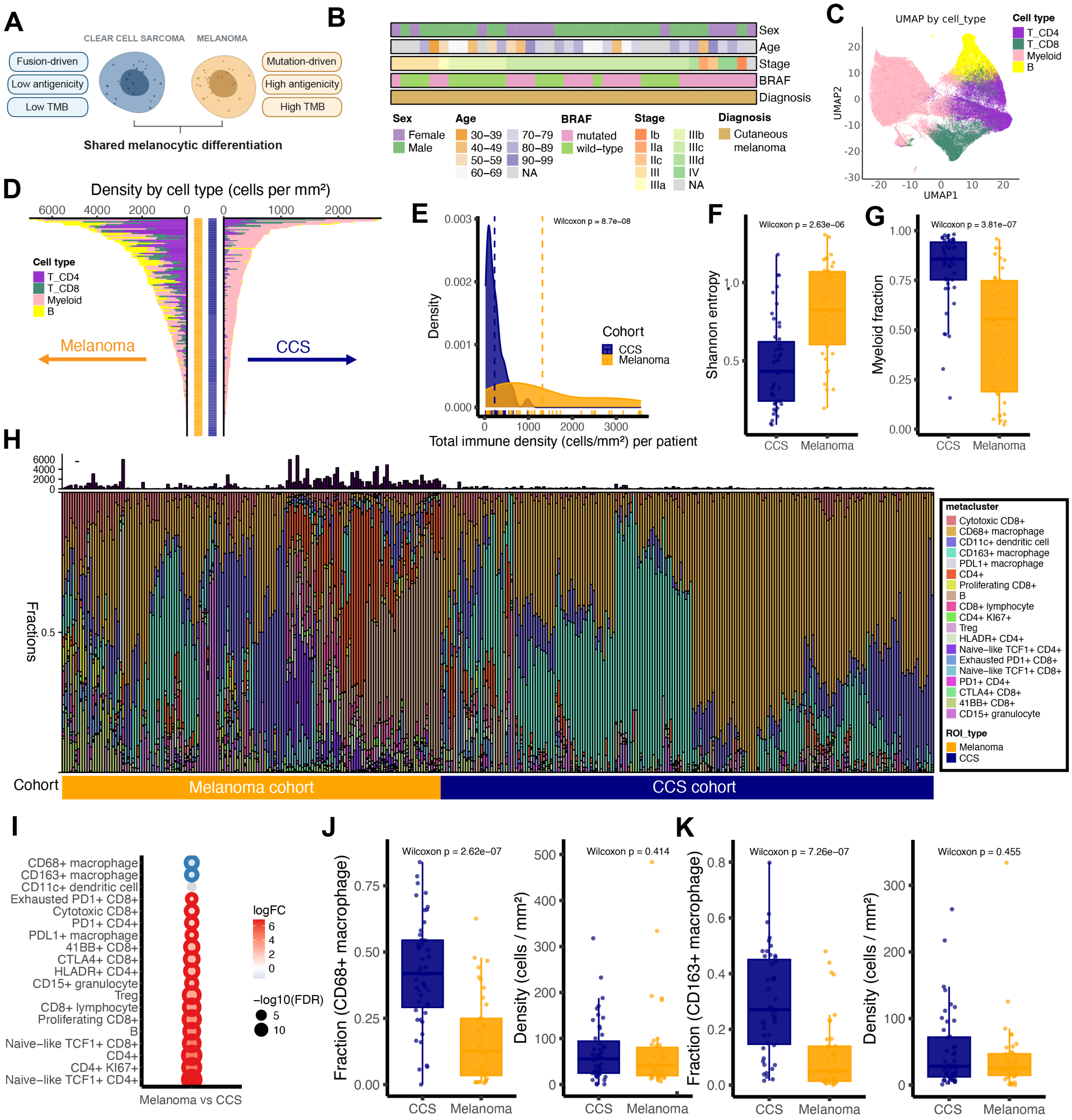


**Figure S7: Comparison of the CCS and cutaneous melanoma immune ecosystems.** *Related to* ***Figure 2****.*

**(A)** Schematic of the two entities: CCS (fusion-driven, low antigenicity, low tumour mutational burden) and cutaneous melanoma (driven by driver-gene mutations, high antigenicity, high tumour mutational burden), with shared melanocytic differentiation (created with BioRender.com).

**(B)** Metadata of the profiled cutaneous-melanoma patients (152 ROIs from 38 patients).

**(C)** UMAP of the single immune cells of the combined CCS and cutaneous-melanoma cohorts (10% downsampled for visualization), coloured by broad immune cell phenotype.

**(D)** Mirrored bar plots of the immune cell density (cells/mm²) per ROI, split by cohort: cutaneous melanoma (left, orange) and CCS (right, blue). Within each ROI, bars are partitioned by the indicated immune cell type and the bar lengths sum to the total immune cell density of the ROI.

**(E)** Distribution of the total immune density per patient (kernel density estimate) for CCS (blue) and cutaneous melanoma (orange); dashed vertical lines mark the cohort medians. P values are indicated (Wilcoxon rank-sum test).
**(F)** Box plots of the Shannon entropy of the broad immune cell types per patient in CCS (blue) and cutaneous melanoma (orange). Box plots show the median, interquartile range (IQR), and whiskers extend to the most extreme values within 1.5×IQR from the quartiles. P values are indicated (Wilcoxon rank-sum test).
**(G)** Box plots of the myeloid fraction (out of all immune cells) per patient in CCS (blue) and cutaneous melanoma (orange), drawn as in F. P values are indicated (Wilcoxon rank-sum test).
**(H)** Stacked bar plots of the fractional composition of the indicated immune subtypes per ROI; the histogram above shows the absolute immune cell count per ROI.
**(I)** Differential abundance by edgeR quasi-likelihood F-test of the immune subtypes between cutaneous melanoma and CCS. Dot size encodes −log10(FDR), colour encodes log2 fold change (cutaneous melanoma vs CCS), and the ring marks cell types called at a false discovery rate below 0.1 with BH correction.
**(J)** Box plots of the fraction (left) and the density (right) of $CD68^+$ macrophages in CCS (blue) and cutaneous melanoma (orange), drawn as in F. P values are indicated (Wilcoxon rank-sum test).
**(K)** As in J, for $CD163^+$ macrophages.

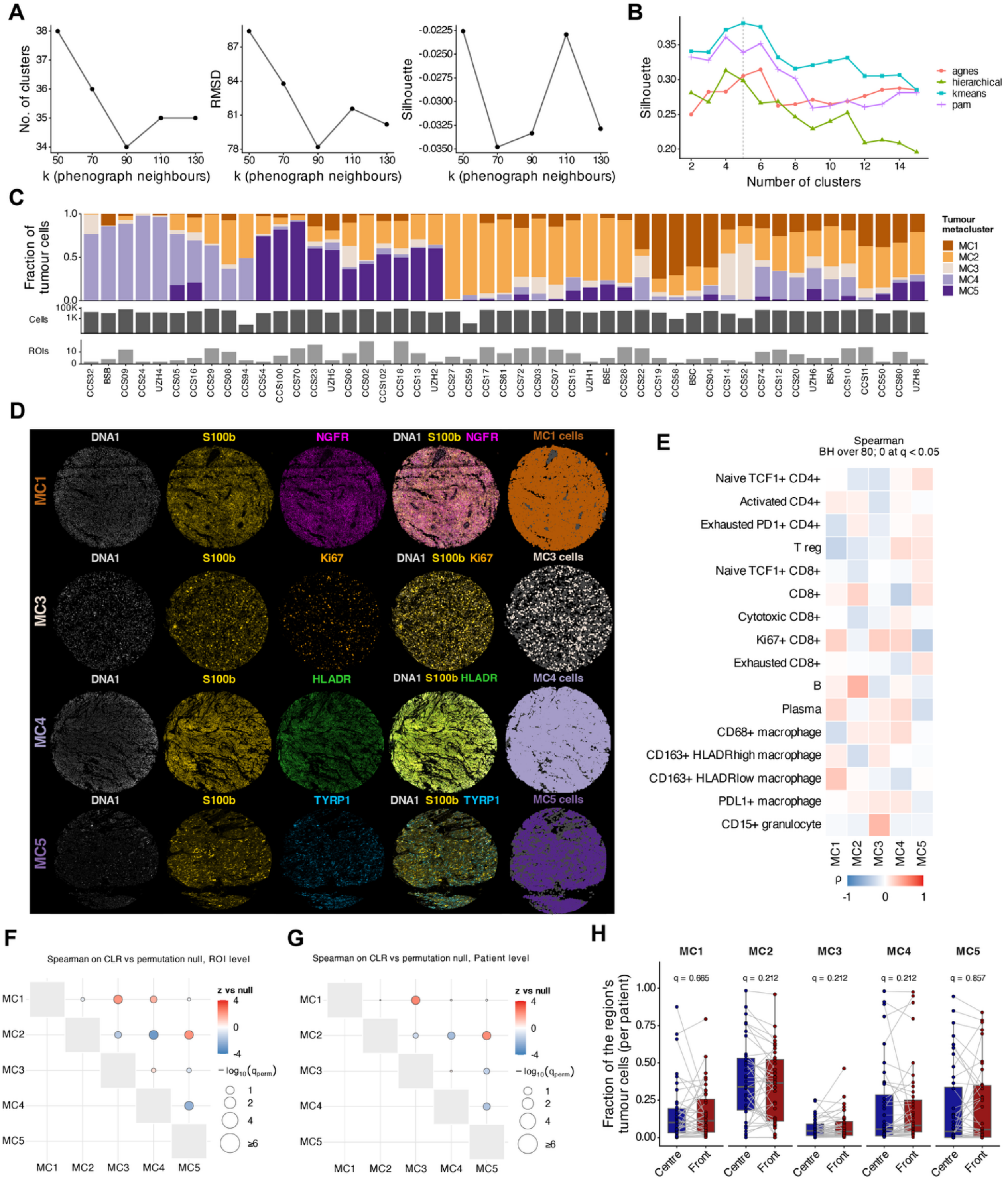


**Figure S8: Tumour metaclusters: clustering, composition, regional distribution and co-occurrence.** *Related to* ***Figure 3.***

**(A)** Parameter selection for graph-based clustering of tumour cells. The number of clusters (left), the root-mean-square deviation (RMSD; centre) and the silhouette width (right) are plotted against the PhenoGraph neighbourhood size k, the number of nearest neighbours used to build the cell graph. A k of 110 was selected for downstream analysis.

**(B)** Silhouette width for different numbers of tumour-cell clusters and four algorithms (hierarchical, agnes, k-means and PAM). Each grouping is scored by its silhouette width. The dashed line marks five clusters, the number of metaclusters used.
**(C)** Stacked bar plots of the per-patient tumour metacluster composition; the bar plots below indicate the number of cells and the number of ROIs per patient.
**(D)** Representative IMC single-marker and composite images of the tumour cell metaclusters, with the segmented cells assigned to each metacluster at right. MC2, which has no defining marker in the panel, is not shown.
**(E)** Heatmap of the Spearman correlation between each tumour metacluster and each of 16 immune subtypes, computed per patient on centred log-ratio-transformed fractions; Benjamini-Hochberg correction over the 80 pairs tested (5 metaclusters × 16 immune subtypes).
**(F, G)** Correlations between tumour metacluster fractions at the ROI (F) and patient (G) levels. Spearman correlations of centred log-ratio-transformed fractions were compared with a permutation null accounting for compositional constraints. Colour indicates the z-score relative to the null; dot size indicates $-\log10(q)$ with Benjamini-Hochberg correction.
**(H)** Box plots of the per-ROI fraction of the tumour cells of each metacluster, shown for the tumour centre versus invasive front. Box plots show the median, interquartile range (IQR), and whiskers extend to the most extreme values within 1.5×IQR from the quartiles; each point is one patient, and lines connect the two regions of a patient. BH-adjusted q values over the five metaclusters are indicated (paired Wilcoxon signed-rank tests).

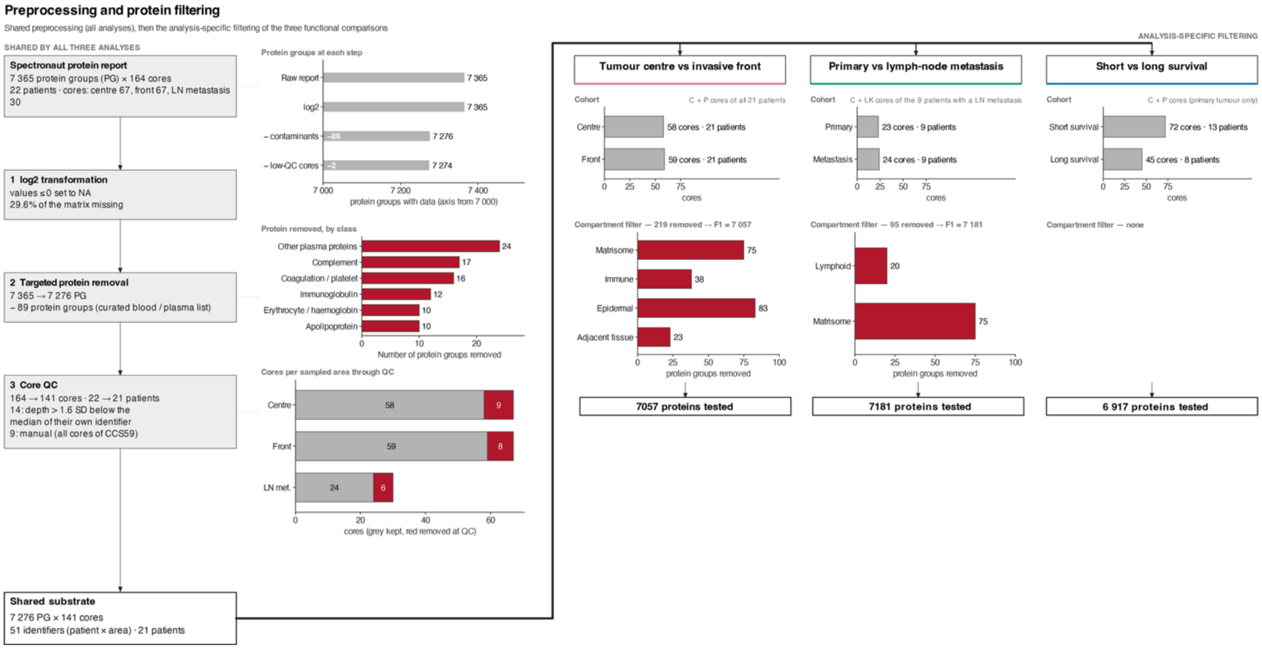


**Figure S9: Deep visual proteomics preprocessing and protein filtering.** *Related to* ***Figure 4***.

Flowchart from the Spectronaut protein report for the proteins tested in each of the three functional comparisons (i.e., tumour centre versus invasive front, primary tumour versus lymph-node metastasis, and short versus long survival). The shared preprocessing is shown at left. The analysis-specific compartment filtering for the three functional comparisons is shown at right. The bar plots show the number of proteins at each step.

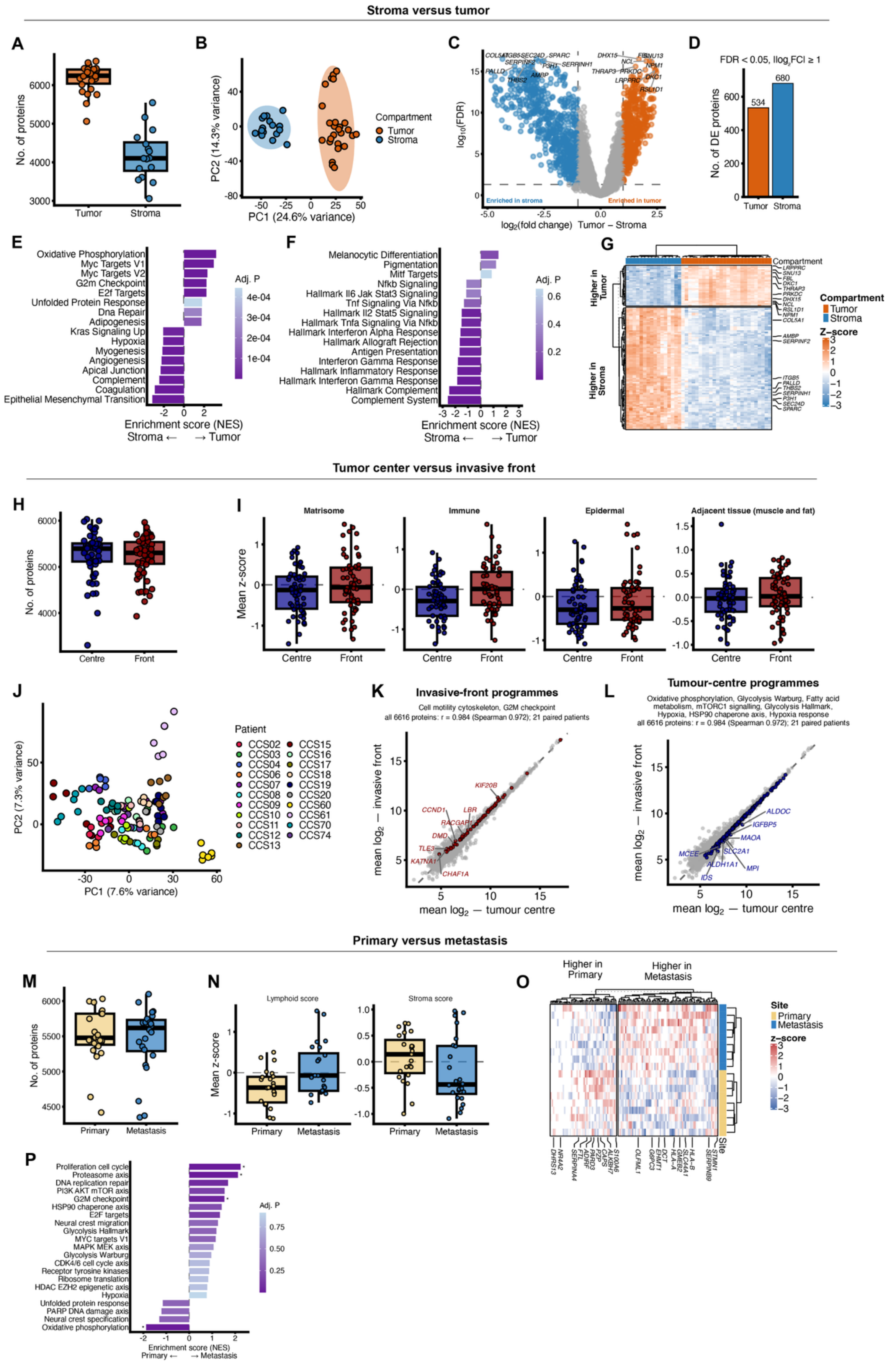

Stroma versus tumor
A
No. of proteins
Tumor
Stroma
B
PC2 (14.3% variance)
PC1 (24.6% variance)
Compartment
Tumor
Stroma
C
log10(FDR)
log2(fold change) Tumor – Stroma
Enriched in stroma
Enriched in tumor
D
FDR < 0.05, |log2FC| ≥ 1
No. of DE proteins
534
680
Tumor
Stroma
E
Oxidative Phosphorylation
Myc Targets V1
Myc Targets V2
G2m Checkpoint
E2f Targets
Unfolded Protein Response
Dna Repair
Adipogenesis
Kras Signaling Up
Hypoxia
Myogenesis
Angiogenesis
Apical Junction
Complement
Coagulation
Epithelial Mesenchymal Transition
Adj. P
Enrichment score (NES)
Stroma ← → Tumor
F
Melanocytic Differentiation
Pigmentation
Mitf Targets
Nfkb Signaling
Hallmark Il6 Jak Stat3 Signaling
Tnf Signaling Via Nfkb
Hallmark Il2 Stat5 Signaling
Hallmark Tnfa Signaling Via Nfkb
Hallmark Interferon Alpha Response
Hallmark Allograft Rejection
Antigen Presentation
Interferon Gamma Response
Hallmark Inflammatory Response
Hallmark Interferon Gamma Response
Hallmark Complement
Complement System
Adj. P
Enrichment score (NES)
Stroma ← → Tumor
G
Compartment
Higher in Tumor
Higher in Stroma
Compartment
Tumor
Stroma
Z–score
Tumor center versus invasive front
H
No. of proteins
Centre
Front
I
Matrisome
Immune
Epidermal
Adjacent tissue (muscle and fat)
Mean z-score
Centre
Front
J
PC2 (7.3% variance)
PC1 (7.6% variance)
Patient
CCS02
CCS03
CCS04
CCS06
CCS07
CCS08
CCS09
CCS10
CCS11
CCS12
CCS13
CCS15
CCS16
CCS17
CCS18
CCS19
CCS20
CCS60
CCS61
CCS70
CCS74
K
Invasive-front programmes
Cell motility cytoskeleton, G2M checkpoint
all 6616 proteins: r = 0.984 (Spearman 0.972); 21 paired patients
mean log2 — invasive front
mean log2 — tumour centre
L
Tumour-centre programmes
Oxidative phosphorylation, Glycolysis Warburg, Fatty acid metabolism, mTORC1 signalling, Glycolysis Hallmark, Hypoxia, HSP90 chaperone axis, Hypoxia response
all 6616 proteins: r = 0.984 (Spearman 0.972); 21 paired patients
mean log2 — invasive front
mean log2 — tumour centre
Primary versus metastasis
M
No. of proteins
Primary
Metastasis
N
Lymphoid score
Stroma score
Mean z-score
Primary
Metastasis
O
Higher in Primary
Higher in Metastasis
Site
Primary
Metastasis
z–score
P
Proliferation cell cycle
Proteasome axis
DNA replication repair
PI3K AKT mTOR axis
G2M checkpoint
HSP90 chaperone axis
E2F targets
Neural crest migration
Glycolysis Hallmark
MYC targets V1
MAPK MEK axis
Glycolysis Warburg
CDK4/6 cell cycle axis
Receptor tyrosine kinases
Ribosome translation
HDAC EZH2 epigenetic axis
Hypoxia
Unfolded protein response
PARP DNA damage axis
Neural crest specification
Oxidative phosphorylation
Adj. P
Enrichment score (NES)
Primary ← → Metastasis

**Figure S10: Deep visual proteomics comparing stroma against tumour, tumour centre against invasive front, and primary tumour against lymph-node metastasis.** *Related to **Figures 4 and 6**.*

**(A–G)** Comparison of stroma against tumour in a separate pilot acquisition (42 cores from 4 patients: 26 tumour, 16 stroma).

**(A)** Box plot of the number of proteins identified per core, in tumour and stroma. Box plots show the median, interquartile range (IQR), and whiskers extend to the most extreme values within 1.5×IQR from the quartiles; points denote individual cores.

**(B)** Principal component analysis of the 42 cores, coloured by compartment, with the 95% confidence ellipse per compartment.

**(C)** Volcano plot of differential protein expression, tumour versus stroma. Differential abundance was tested with a moderated t test (limma). Proteins called at a false discovery rate below 0.05 and an absolute log2 fold change of at least 1 are coloured and labelled. Benjamini-Hochberg correction over all tested proteins.

**(D)** Bar plot of the number of differentially abundant proteins in C.

**(E, F)** Gene-set enrichment (fgsea) of all quantified proteins, ranked by the moderated t statistic for tumour versus stroma, against the Hallmark collection (E) and curated pathway sets (F); bars point towards the compartment in which each set is enriched. Bars show the normalized enrichment score (NES) and colour denotes the adjusted P value.

**(G)** Hierarchical clustering of the 120 top differential proteins (z-scored; one column per core), split by compartment.

**(H–L)** Comparison of tumour centre against invasive front (117 cores from 21 patients).

**(H)** Box plot of the number of proteins identified per core in each indicated region.

**(I)** Box plots of the mean z-score per core of four curated compartment gene sets (matrisome, immune, epidermal, and adjacent tissue: muscle and fat), used to check how much non-tumour tissue each region contains.

**(J)** Principal component analysis of the protein profiles from all analysed cores (n = 117), coloured by patient.

**(K, L)** Scatter plots of protein abundance at the tumour centre (x) against the invasive front (y) for the 21 patients with samples from both regions. Each point is one protein (6,616 proteins). Proteins are coloured by the leading-edge proteins (the proteins that drive the enrichment score in the gene-set enrichment analysis of Figure 4C) of the programmes enriched at the invasive front (K; cell motility/cytoskeleton and G2/M checkpoint) or in the tumour centre (L; oxidative phosphorylation, Glycolysis Warburg, fatty-acid metabolism, mTORC1 signalling, Glycolysis Hallmark, hypoxia, the HSP90 chaperone axis and hypoxia response).

**(M–P)** Comparison of primary tumour against lymph-node metastasis (47 cores from 9 patients).

**(M)** Box plot of the number of proteins per core in primary versus metastatic samples.

**(N)** Box plots of the mean lymphoid score (left) and stroma score (right) per core, in primary versus metastatic samples.

**(O)** Hierarchical clustering of the 120 top differential proteins (z-scored; one column per core), split by site.

**(P)** Gene-set enrichment (fgsea) of all quantified proteins, ranked by the moderated t statistic for primary tumour against lymph-node metastasis, against curated pathway sets; bars point towards the site in which each set is enriched. Bars show the normalized enrichment score (NES) and colour denotes the adjusted P value.

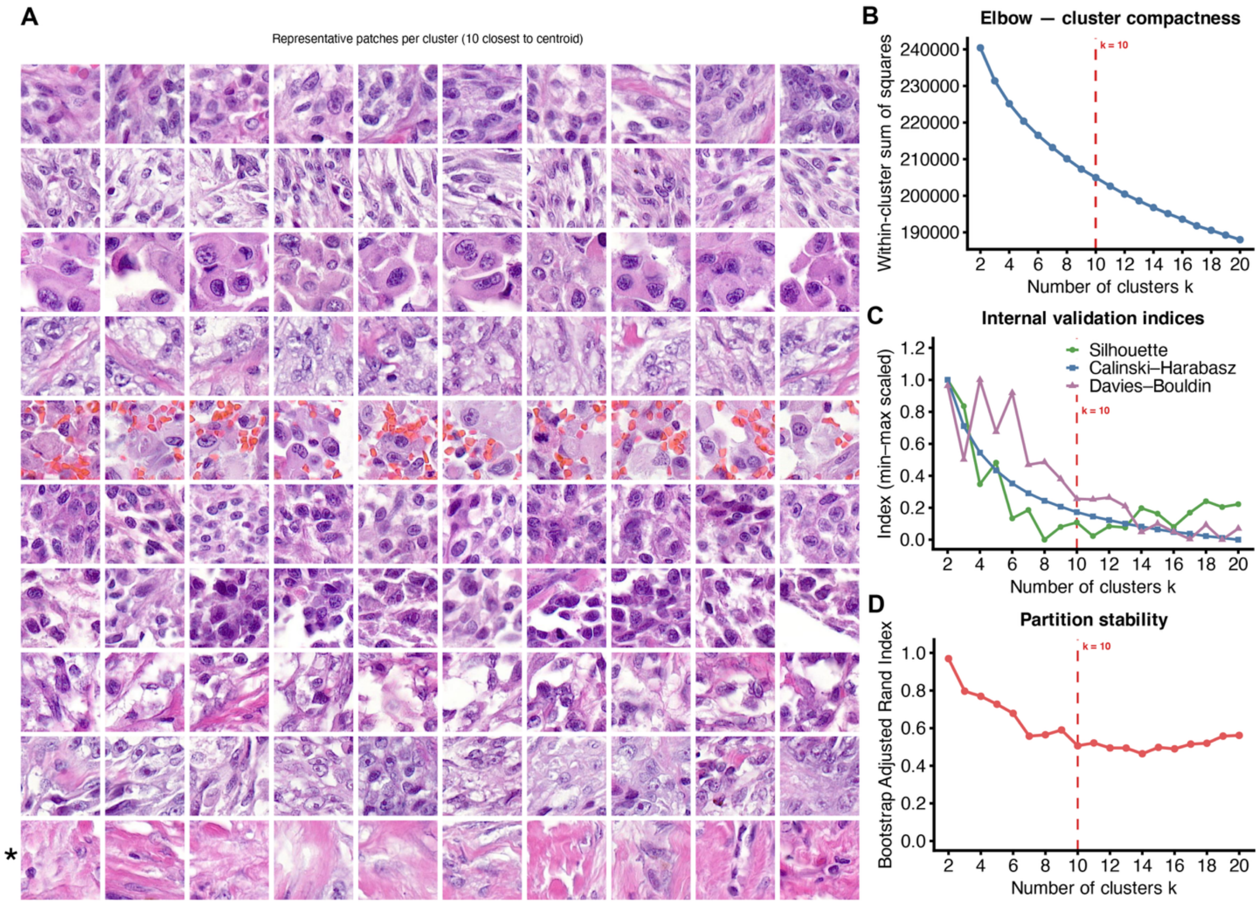


**Figure S11: H&E patch clustering.** *Related to **Figure 5**.*

**(A)** H&E images of representative patches per cluster for each of the 10 k-means clusters of the H&E patch features. In each case, the 10 patches closest to the cluster centroid are shown. The cluster marked with an asterisk was annotated as non-tumour by pathologist review.

**(B)** Within-cluster sum of squares against the number of clusters k (k = 2–20), for clustering of H&E patch features; the dashed line marks the chosen k = 10 (combining pathologist review).

**(C)** Silhouette, Calinski-Harabasz and Davies-Bouldin indices, each min-max scaled over k = 2–20. Higher values are better for the silhouette and Calinski-Harabasz indices, lower values for the Davies-Bouldin index; the dashed line marks the chosen k = 10 (combining pathologist review).

**(D)** Partition stability: for each k, k-means was refitted on 15 bootstrap resamples of the patch features, and the agreement of each refit with the partition of the full data was measured by the adjusted Rand index; the mean over the 15 refits is plotted, and the dashed line marks k = 10.
